# Temperature and the Composition of Diets: Evidence from China

**DOI:** 10.1101/2025.04.08.25325375

**Authors:** Xi Chen, Shuo Li, Ding Ma, Jintao Xu

## Abstract

How climate change affects human welfare depends not only on what food systems produce but on what individuals eat once food is on the table. Using three-day dietary recalls from the China Health and Nutrition Survey matched to county weather in the same three-day window, we show that short-run departures from comfort temperature raise the fat share of energy intake and the probability of a diet whose fat share exceeds the WHO threshold. Heat and cold raise the fat share through opposite margins, though the cold response is smaller and less precisely estimated. Under heat, total energy, carbohydrate, and protein intake fall, while absolute fat intake does not change detectably, so the fat share rises as a composition effect, even as total calories decline. Under cold, absolute fat intake appears to rise more than other macronutrients, an intake effect. A macronutrient decomposition is therefore essential: identical movements in total calories mask very different movements in diet quality, a distinction that calorie-or index-based measures cannot deliver. The responses align with physiological appetite regulation, though our design cannot separate physiology from other demand-side channels. Effects are larger for rural and lower-income households but smaller for those that own cooling and heating appliances. We read individual diet as a behavioral input to health capital through which temperature, and adaptive capacity, shape nutritional inequality.

## 1. Introduction

How much does climate change cost human welfare, and through which channels? Economists have built this accounting from several well-studied margins: crop yields and agricultural productivity (Deschênes and Greenstone, 2007; Burke and Emerick, 2016; Aragón et al., 2021), mortality and morbidity (Deschênes and Greenstone, 2011; Gasparrini et al., 2015; Carleton et al., 2022), labor supply and time use (Graff Zivin and Neidell, 2014), and energy demand and adaptation (Barreca et al., 2016). A common feature of this literature is that temperature affects welfare through what the economy produces and how people work, with food treated mainly as a quantity constrained by supply. What people actually eat once food is on the table has received much less attention, even though realized nutrition is the direct input to health capital (Grossman, 1972). This paper asks whether short-run deviations from comfort temperature change individual dietary composition, and whether the response pushes diets toward the high-fat patterns associated with obesity and cardiometabolic disease (WHO, 2023a; Ng et al., 2025).

Diet is a natural place to look for a missing channel because temperature plausibly moves both the amount and the mix of food that households consume. Heat can suppress appetite and reduce total intake, while carbohydrates and protein generate more diet-induced thermogenesis during digestion than fat does (Westerterp, 2004). Cold can raise energy requirements through shivering and brown-adipose-tissue thermogenesis (Virtanen et al., 2009). Fat delivers 9 kcal per gram, compared with 4 kcal per gram from carbohydrates or protein. These asymmetries imply that heat and cold can both raise the share of energy from fat, but through different adjustment margins. Calorie-based measures can therefore miss a deterioration in diet quality. A diet that loses carbohydrates and protein without losing fat is compositionally riskier, even when total calories fall. Whether these forces are large enough to matter at the frequency of weather, rather than as long-run climate, is an empirical question.

We answer this question using the China Health and Nutrition Survey, which records individual food intake through three consecutive 24-hour recalls validated against household-level weighing and enumerator-assisted recall by trained nutritionists. The analysis covers 28,941 individuals in 7,057 households from nine provinces between 1991 and 2011. We match each respondent’s three-day recall window to county-level daily weather. The estimating equation relates diet in the recall window to temperature exposure in the same window, conditional on individual, province-by-year, province-by-month, and year-by-month fixed effects and contemporaneous controls for precipitation, humidity, wind, and sunshine. The identifying variation is deliberately short-run and within-individual. It compares the same person across survey waves that happen to fall under different three-day temperature conditions, and it compares respondents who live in the same province-month cell but are interviewed on different dates. It is not driven by long-run differences in climate across places, by long-run trends in diet over time, or by seasonal patterns shared across a province.^1^

Our analysis reveals that short-run temperature deviations around the comfort range shift dietary composition toward higher fat share and increase the probability of a high-fat diet, as defined by the World Health Organization’s threshold of 30% of total energy intake (WHO, 2023b). These compositional shifts carry potential health consequences, as high-fat diets are closely linked to obesity (Hariri and Thibault, 2010), promote insulin resistance (Kumar et al., 2021), accelerate atherosclerosis (Lavillegrand et al., 2024), and even contribute to cancer progression (Chen et al., 2024). The macronutrient decomposition reveals that the two channels operate differently. High temperatures significantly reduce the intake of carbohydrates and proteins while leaving fat consumption relatively unaffected, mechanically elevating fat’s share of remaining energy intake; low temperatures elevate overall caloric intake with the largest gains concentrated in fat, a pattern consistent with elevated thermogenic energy demand, though the cold-side estimates are less precise.

We discuss the mechanisms underlying these dietary responses. The observed patterns align with physiological pathways in biomedical literature: appetite suppression under heat and elevated energy demand under cold. Because both heat and cold reduce working hours, the dietary shift is not driven by higher activity-related energy needs. The evidence points to physiological appetite regulation, though our design cannot separate it from intake adjustments due to lower energy requirements or other demand-side channels.

Furthermore, our analysis identifies significant socioeconomic disparities in dietary responses. Rural and lower-income households are disproportionately sensitive to temperature-induced dietary changes, reflecting structural disadvantages including limited food storage infrastructure, constrained market access, and lower ownership of cooling and heating appliances. Indeed, appliance ownership represents one plausible channel through which socioeconomic advantages translate into dietary resilience. These disparities carry long-term implications for human capital development, productivity, and economic inequality (Maluccio et al., 2009; Almond et al., 2015; Chong et al., 2016; Hjort et al., 2017; Coffey et al., 2018), and point to the importance of targeted adaptation policies for vulnerable populations.

This study contributes to the literature in four ways. First, we add an individual dietary-demand margin to research on climate change, food systems, and nutrition. Studies in this area have characterized how weather and climate shift crop yields, agricultural productivity, and food availability (Deschênes and Greenstone, 2007; Burke and Emerick, 2016; Aragón et al., 2021), and how these changes translate into household food acquisition and food security (Stainier et al., 2025).^2^ These supply-side channels determine what food can reach the table. We study a complementary demand-side channel: how individuals change macronutrient composition once food is on the table. Matching three-day dietary recalls to the same three-day weather window isolates a contemporaneous, high-frequency dietary response that growing-season and annual weather measures cannot capture, and that aggregate food-security outcomes do not decompose.

Second, relative to the climate change and health literature, we extend the set of health-relevant outcomes from direct mortality and morbidity to the behavioral input that shapes chronic disease risk. Existing work documents temperature effects on mortality, morbidity, mental health, labor supply, time use, and average body weight (Deschênes and Greenstone, 2011; Graff Zivin and Neidell, 2014; Gasparrini et al., 2015; Agarwal et al., 2021; Ebi et al., 2021; Huang and Hong, 2024), and Churchill and Srivastava (2025) show that short-run temperature alters diet quality indices and weight-management efforts in the United States. We decompose the dietary response into three macronutrients in a developing-country setting and show that heat and cold worsen diet quality through different adjustment margins. The decomposition implies that identical changes in total calories can hide very different movements in fat share, and it provides an empirical input for mapping repeated weather shocks into cumulative diet quality. Our design does not estimate long-run disease outcomes directly; instead, it measures a short-run margin on which those outcomes depend.

Third, we contribute to the emerging literature on environmental determinants of diet. Recent studies show that air pollution shifts household food purchases away from unhealthy products toward healthier ones, a response that operates through avoidance (Agarwal et al., 2025; Huang et al., 2025). Temperature differs from pollution along two dimensions that make it a distinct, and in some respects more informative, environmental stressor. One, it is bidirectional, so the same outcome can be studied at both tails of the exposure distribution; we exploit this to show that heat and cold worsen diet quality through opposite adjustment margins, a composition effect under heat and an absolute-intake effect under cold, which a one-sided stressor cannot reveal. Two, it acts on the body rather than only on behavior around the body; the responses we document are consistent with appetite suppression and diet-induced thermogenesis under heat and with elevated thermoregulatory energy demand under cold, so the dietary response is not simply a by-product of avoidance, although our design cannot rule out other demand-side channels.

Finally, our setting is a developing country in the midst of a nutrition transition, with the sample’s high-fat-diet prevalence rising from 22% to 57% between 1991 and 2011. Complementing well-documented temperature effects on nutrition-transition in developed countries (e.g., Churchill and Srivastava, 2025), we show that, in a population moving toward the WHO fat threshold from below, off-comfort temperatures push diets across it. Environmental stressors thus enter nutrition not only through food supply but through individual intake, and they do so unequally.

## 2. Conceptual Framework

Our framework models macronutrient choice as the outcome of a temperature-dependent preference over carbohydrate, protein, and fat, subject to an energy requirement, a physical intake limit, and a budget constraint. Temperature influences selection through two primary physiological channels. First, the digestion of carbohydrate and protein releases more heat than that of fat (Westerterp, 2004). This metabolic heat is beneficial in cold environments but aversive in the heat; consequently, the appeal of carbohydrate and protein relative to fat declines as temperatures rise. Second, cold exposure raises the body’s energy requirement through shivering and brown-adipose-tissue thermogenesis (Virtanen et al., 2009). Because fat provides at 9 kcal per gram compared to 4 for the other two macronutrients, it represents the most efficient energy source to meet this increased demand. Conversely, heat suppresses appetite and lowers overall energy requirements.

The two physiological channels imply the same directional change for the fat share of energy, but through distinct behavioral anatomy. Under heat, carbohydrate and protein intake fall while fat intake changes little, so the fat share rises because the total energy denominator shrinks. Under cold, fat intake rises more sharply than the other macronutrients, meaning the fat share rises because the fat-specific numerator grows. The framework does not a priori determine which channel dominates at a given temperature. Instead, it identifies two candidate margins and the macronutrient signatures that distinguish them, which are tested in Section 5. Appendix A provides a formal model.

## 3. Data

Our analysis draws on two primary data sources: household survey data and high-resolution historical weather information.

### 3.1 Household data

We use the individual-level data from the China Health and Nutrition Survey (CHNS). It encompasses 28,941 individuals from 7,057 households in 9 provinces from 1991 to 2011 (see Figure A1(a)).^3^ This dataset has three advantages. First, it provides detailed individual-level information on nutrition. Such data are rarely available: purchase records do not reveal actual consumption, and large-scale longitudinal surveys with dietary intake measures are uncommon. The CHNS overcomes this limitation by offering repeated, precise records of food and nutrient intake, enabling us to directly observe what people consume rather than inferring it indirectly (for the questionnaire on food consumption, see Figure A2). In addition, it contains information on demographics, socioeconomic status, and health outcomes. Second, its nine provinces span diverse climatic zones. Finally, the timing of the CHNS surveys, primarily conducted between September and November (see Figure A1(b)), offers distinct empirical advantages. On the one hand, temperatures during these months are relatively moderate yet still sufficiently variable, providing an ideal setting to observe sudden hot or cold shocks rather than prolonged, stable extreme temperatures. On the other hand, this period falls after the main growing season, when yields are already determined. Thus, the potential confounding effects of fluctuations in crop growth can be substantially mitigated.

Following the *Chinese Dietary Reference Intakes—Macronutrient (WS/T 578.1-2017)*, we calculate energy intake from carbohydrate, protein, and fat at 4, 4, and 9 kcal per gram, respectively. For the data cleaning process, see Appendix B.

### 3.2 Meteorological data

The meteorological data come from the China Meteorological Data Sharing Service (CMDSS) system. It includes daily average temperature, wind speed, accumulated rainfall, relative humidity, and sunshine hours from more than 2000 weather stations in China. We construct the daily county-level weather variables by taking an inverse-distance-squared weighted average of all the stations located within about an 80 km (50-mile) radius of the county centroid.^4^

Because dietary intake in the CHNS is measured over three consecutive days, we construct temperature exposure within the same three-day window. To capture the intensity of heat and cold during this period, we use cumulative cooling degree days (CDD) and heating degree days (HDD). This approach provides a more precise characterization of short-term thermal exposure than conventional temperature bins, since CDD and HDD measure not only whether the daily average temperature crosses a threshold but also by how much it exceeds or falls short of it. Given the respondents’ average daily temperature distribution (in Figure A1(c)), we choose 25°C and 5°C as the thresholds, which may not appear particularly extreme but are intended to capture meaningful deviations from the thermal comfort range.^5,6^

### 3.3 Summary statistics

Table A2 shows the summary statistics of the variables used in this paper, providing two important messages. First, the overall 3-day energy intake for the entire sample was approximately 6398 kcal, with a protein intake of around 195 g (about 2,130 kcal and 65 g/day). According to the *Chinese Dietary Reference Intakes—Macronutrient (WS/T 578.1-2017)*, adults’ recommended daily energy intake ranges from approximately 2100 to 3200 kcal, while the reference protein intake is about 55-65 g/day. Considering the inclusion of children in the sample, the average values for the entire sample may exhibit slight deviations from those of the population aged 18 years and older. Figure A5 highlights two notable trends regarding dietary fat consumption from 1991 to 2011 in our sample. The percentage of individuals consuming a high-fat diet (defined as more than 30% of dietary energy coming from fat) (Wang et al., 2020) exhibited a rapid increase from 22.44% in 1991 to 56.95% in 2011. Meanwhile, the average percentage of total dietary energy derived from fats also steadily increased from 22.06% in 1991 to 32.38% in 2011. This upward trajectory underscores a clear dietary shift in China toward greater fat consumption during the study period, reflecting both nutritional transition and increasing risk factors associated with obesity and other non-communicable diseases.

Second, the distributions of 3-day CDD and HDD are broadly comparable, with mean values of 0.668 and 0.504 degree-days, respectively. This is largely shaped by survey timing. As shown in Figure A1(b), most CHNS surveys were conducted between September and November. Consequently, respondents were typically exposed to average temperatures between 5 and 25°C, with relatively few observations below 0°C or above 30°C (see Figure A1(c)). The chosen cutoffs of 5°C and 25°C are approximately at the 5th and 95th percentiles of the 3-day average temperature.

## 4. Empirical Strategy

We estimate the effects of off-comfort temperatures on diet using ordinary least squares (OLS). Because short-term temperature fluctuations are beyond individual control, they can be regarded as quasi-random once rich sets of fixed effects are included. Moreover, the CHNS dietary surveys were conducted at varying dates within each year, creating additional quasi-random variation in temperature exposure even among individuals living in the same county. We examine this assumption directly with a balance test of interview timing (Appendix C). Predetermined characteristics are not perfectly balanced across the temperature distribution, but the estimates are unchanged when these characteristics are controlled for. We estimate the following regression:

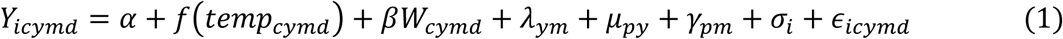

where *Y_icymd_* represents the outcome variables over the past three days for individual *i* in county *c*, year *y*, month *m*, and day *d*. First, we focus on dietary composition, measured by the proportion of energy from fat and the high-fat diet index, as our primary outcomes. Second, to understand the channels through which dietary composition shifts, we examine overall food consumption and total energy intake. Third, we further decompose macronutrient intake into carbohydrates, proteins, and fats. For the proportion of energy from fat and the high-fat diet index, we use the actual value, as they are already measured in percentage terms. For total consumption, energy, carbohydrates, proteins, and fat, we use the logarithmic form to capture proportional changes.

The key independent variable is *temp_cymd_* , which denotes the daily average temperature. Its functional form, *f*(*temp_cymd_*) , captures the flexible measure of temperature. In our baseline specification, we employ the cumulative CDD (above 25°C) and HDD (below 5°C) over the past three days to measure short-term exposure to high and low temperatures, respectively. Alternative definitions of temperature exposure are explored in the robustness checks. *W_cymd_* is a vector of the 3-day average meteorological variables other than temperature, including precipitation, relative humidity, wind speed, and sunshine hours.

To address potential confounding factors, we incorporate a rich set of fixed effects at multiple levels. Year-month fixed effects (*λ_ym_*) control for nationwide changes, such as economic development that expands food availability and quality, and climate trends. Province-year fixed effects (*μ_py_*) absorb province-specific variation across years in growing season heat accumulation, which helps remove potential confounding from the agricultural supply side. Province-month fixed effects (*γ_pm_*) capture seasonal and regional differences in food supply and consumption habits. For instance, local harvest cycles and dietary traditions vary across provinces, and typical seasonal temperatures also differ systematically between the north and the south. With these fixed effects, we exploit only deviations from the usual seasonal climate within the same province.^7^ Individual fixed effects (*σ_i_*) absorb all time-invariant personal characteristics, such as baseline health status, inherent dietary preferences, or cultural eating habits. These fixed effects purge systematic variation from both the temperature and dietary sides, leaving short-term fluctuations in temperature plausibly random. The error term (*ε_icymd_*) is clustered at the individual level to account for within-person correlation over time.

A separate identification concern is that temperature may affect dietary outcomes through agricultural supply rather than demand. Heat or cold could reduce local crop yields, tighten food availability, and raise retail prices, leading households to substitute across macronutrients in ways that mimic a physiological response. We address this concern in two complementary ways. First, our identifying variation is inherently ill-suited to capture supply-side effects. The CHNS surveys were conducted at varying dates within each year, and our temperature measures reflect idiosyncratic three-day fluctuations tied to the exact timing of each household’s interview. Such short-run deviations are far too transient to affect seasonal crop yields or propagate through the agricultural supply chain to retail markets. Supply-side confounding would require that a randomly timed three-day temperature shock during the survey window coincide with measurable local price or availability changes, which is implausible given the multi-month lag^8^ between field-level weather shocks and retail price adjustments. Second, China’s grain reserve and price stabilization system provides an additional institutional buffer.^9^ This institutional architecture is designed precisely to prevent short-run weather events from translating into retail price volatility, further attenuating any supply-side channel within our observation window^10^.

A potential concern with nonlinear temperature specifications is that warming-induced trends in off-comfort temperature exposure may correlate with place-specific trends in outcome variables. This correlation can generate spurious effects even in panel regressions, a bias documented by Jones et al. (2026). Several features of our design mitigate this concern. First, our temperature measures capture idiosyncratic short-run deviations in temperature exposure within a narrowly defined survey window, rather than annual counts of off-comfort temperature days that are more susceptible to trend contamination. The identifying variation is thus driven by quasi-random differences in the exact timing of survey visits across individuals within the same county and year, which is unlikely to be systematically correlated with long-run dietary trends. Second, province-year fixed effects (*μ_py_*) absorb any province-level correlation between long-run temperature trends and dietary trends. Nonetheless, following Jones et al. (2026), we construct counterfactual 3-day CDD and HDD based on the estimated trend component of county-month temperature and include these as additional controls in robustness tests.

## 5. Results

### 5.1 Baseline results

Figure 1(a) (and Table A3) presents the effects of short-term temperature fluctuations on dietary composition and macronutrient intake estimated by equation (1). Both heat and cold stress shift dietary composition toward a higher fat share of energy, although the cold-side estimates are smaller and less precisely estimated. Each additional degree-day above 25°C over the past three days raises the proportion of energy from fat by 0.081 percentage points and increases the likelihood of a high-fat diet by 0.404 percentage points. Each additional degree-day below 5°C increases the fat share by 0.038 percentage points and the probability of a high-fat diet by 0.195 percentage points.^11^ To benchmark these magnitudes, the fat share of energy in our sample rose by about 0.52 percentage points per year from 1991 to 2011 (Figure A5); a single cooling degree-day therefore moves the fat share by roughly one-sixth of a year’s secular trend, so that about six cooling degree-days are equivalent to one full year of the long-run nutrition transition, though this comparison benchmarks the magnitude of a transitory three-day shock against a permanent trend rather than any cumulative exposure.

**Figure 1.**
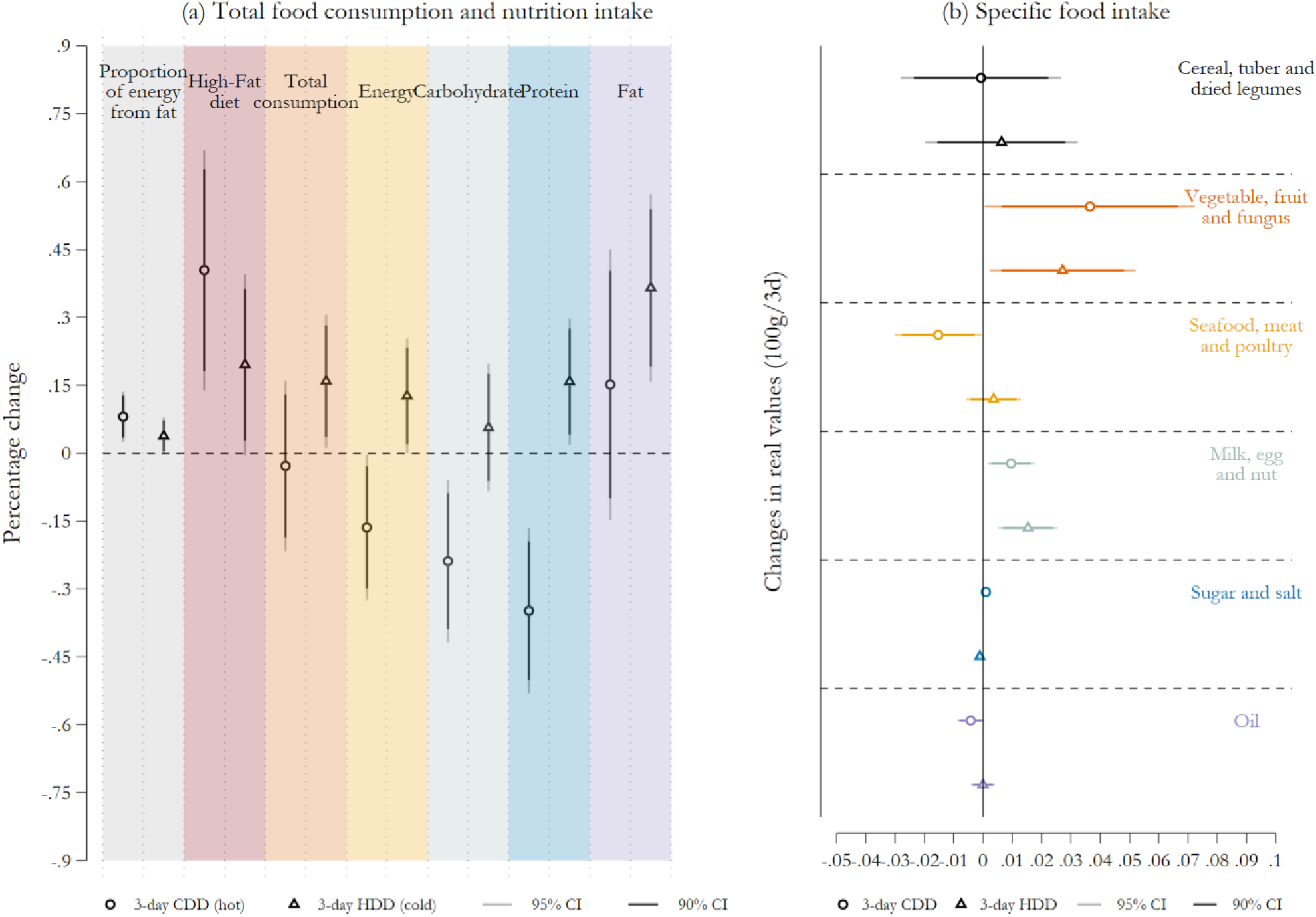
**Temperature affects food consumption and macronutrient intake.***Notes:* These figures illustrate the effects of temperature on 3-day food consumption, macronutrient intake, the proportion of energy from fat, and the probability of high-fat diets (in (a)) and the intake of six groups of food and condiments (in (b)). The foods include cereals, tubers, dried legumes, vegetables, fruits, fungus, seafood, meat, poultry, milk, eggs, and nuts, while the condiments include sugar, salt, and oils. Total consumption is calculated by summing up all the food consumed by the individual, excluding the condiments, which are only usable after 1997. In Figure (a), the outcome variables are the original levels of the proportion of energy from fat, high-fat diets, and the logarithmic form of the value of food consumption or macronutrient intake. For ease of interpretation, we multiply the regression coefficients and CIs by 100 so that the results can be interpreted as a change in percentage. In Figure (b), the outcome variables are the original levels of the intake of each group of food and condiments. As shown in equation (1), the regression model includes individual fixed effects, year-month fixed effects, province-year fixed effects, and province-month fixed effects. The regression results are reported in Table A3 and Table A8. The circles and triangles represent the regression coefficients, and the dark (light) lines represent 90% (95%) confidence intervals. Standard errors are clustered at the individual level.

These compositional shifts carry direct health implications. Both the World Health Organization (WHO, 2023b) and the 2025-2030 Dietary Guidelines for Americans (USDA and USDHHS, 2026) recommend that total fat intake not exceed 30% of total daily energy intake, as diets exceeding this threshold are associated with higher risks of unhealthy weight gain, obesity, cardiovascular disease, type 2 diabetes, and certain cancers. Critically, the threshold is defined on the share of energy from fat rather than on its absolute amount. Under heat stress, total caloric intake declines, yet the fat share of energy rises, because carbohydrates and proteins are suppressed more sharply than fat. The diet, therefore, becomes compositionally riskier even as total consumption falls, representing a deterioration in dietary quality that is not captured by caloric measures alone. Under cold stress, fat intake rises disproportionately relative to carbohydrates and proteins. In both cases, individuals are pushed toward or further above the 30% threshold, with adverse implications for long-run cardiometabolic health.

We next examine the effects of heat and cold on carbohydrates, proteins, and fat separately. Heat significantly reduces energy intake, with each additional degree-day above 25°C decreasing it by 0.164%. The macronutrient decomposition reveals a pronounced asymmetry in which carbohydrate intake falls by 0.239% and protein intake by 0.348%, while the point estimate for fat is positive (0.151%) but statistically indistinguishable from zero. The differential suppression of carbohydrates and proteins relative to fat mechanically elevates fat’s share of remaining energy intake even as total consumption declines. The fat share therefore rises because its denominator contracts, not because fat intake increases. Cold appears to operate through an opposite channel. Specifically, each additional degree-day below 5°C increases food consumption by 0.159% and energy intake by 0.126%, with fat rising by 0.364%, a magnitude more than twice the increase in protein (0.158%) and over six times that of carbohydrates (0.057%). Wald tests reject equality of the fat coefficient with those on carbohydrate and protein under both heat (p<0.01) and cold (p<0.05), confirming that the fat response is statistically distinct rather than merely differing in significance across separate regressions (see Table A4).

These dietary patterns pose potential health risks, particularly obesity, which is associated with severe diseases such as cardiovascular conditions, diabetes, and cancers (Sung et al., 2019; Soerjomataram and Bray, 2021; Ng et al., 2025). Excess calories convert into body fat when not burned. Even stable calorie intake with a higher fat proportion elevates obesity risk. We use OLS to document the association of Body Mass Index (BMI) with energy intake and dietary structure (see Appendix D). Table A5 provides suggestive evidence on the association between dietary composition and health outcomes. Because this is an in-sample OLS correlation, it may partly reflect selection on age, income, and urbanization rather than a causal effect; we therefore rely primarily on the clinical evidence cited above for the health relevance of a higher fat share and read this table only as corroborative. The results reveal two patterns. First, both calorie intake and fat share are positively and statistically associated with BMI and the probability of being overweight. Second, while calorie intake often receives more attention, dietary fat share is associated with an increase in overweight risk, underscoring the importance of compositional changes in diet beyond total energy intake. These correlations are consistent with the concern that temperature-induced shifts toward higher fat share may carry adverse implications for long-run cardiometabolic health, even though the present design does not directly identify the downstream health consequences of these dietary changes.

However, our empirical strategy identifies the deterioration of short-term dietary composition. Whether these transient shifts translate into persistent increases in BMI ultimately depends on the frequency of such temperature shocks and individuals’ intertemporal substitution behaviors. We validate our findings through multiple robustness checks (see Appendix E and Table A6 to Table A7). First, we employ a more stringent specification using individual and province-year-month fixed effects. The point estimates remain stable, though the HDD effect on fat share loses statistical significance, which is likely a consequence of province-year-month fixed effects absorbing much of the identifying variation in cold exposure. Second, to address potential agricultural supply-side confounding, we control for county-year cumulative growing and killing degree days during local main crop seasons, absorbing yield-relevant temperature variation not captured by province-year fixed effects. Third, following Jones et al. (2026), we include counterfactual temperature measures as additional controls to mitigate potential biases from localized warming trends. Fourth, we add day-of-week fixed effects to distinguish weekdays from weekends. Fifth, we add an indicator for major festival periods to control for holiday-related dietary variation. Sixth, we drop all meteorological controls other than temperature. Finally, we test several alternative definitions of temperature exposure. The results are robust across these checks.

### 5.2 Different food groups

We track dietary structure changes through 12 types of food and 3 types of condiments. We simplify these items into six aggregated groups for cleaner presentation. Each group contains zero values (see Table A2), and the quantities are small. Thus, we use the absolute amounts of consumption for each group as the outcome variable instead of the logarithm form after adding one. Temperature-driven consumption changes for each group are presented in Figure 1(b) and Table A8.

Two clear patterns emerge. First, heat primarily reduces the intake of seafood, meat, and poultry. In contrast, hot weather significantly increases the consumption of vegetables, fruits, and fungi, as well as milk, eggs, and nuts, while its effect on cereals, tubers, and dried legumes is not statistically significant. According to China’s Nutrition of Main Foods (China CDC, 2018), we calculate the approximate proportions of three macronutrients supplied by the four major food groups (see Figure A6). Cereals, tubers, and dried legumes are low in fat (nearly 5g per 100g), while milk, eggs, and nuts have more fat (about 26g per 100g), and oils are the most fat-dense. Notably, edible oil accounts for 50% of total fat intake in China (Wang et al., 2020). Thus, hot weather suppresses protein-rich meat intake while simultaneously increasing the consumption of higher-fat items such as eggs and nuts, which keeps total fat intake roughly stable despite the reduction in cooking oils.

Second, cold exposure induces significant increases in vegetables, fruits, and fungi, as well as milk, eggs, and nuts. In contrast, cereals and meat show little response. Because the strongest increases come from fat-rich sources (milk, eggs, and nuts), cold weather also pushes diets toward higher fat intake, consistent with an elevated thermogenic energy demand.

Taken together, these results show that the main drivers of temperature-induced high-fat diets differ across contexts. Heat operates primarily by suppressing protein-rich meats and increasing high-fat animal and plant products, whereas cold stimulates broader demand for fat-rich items. These heat-induced shifts reallocate the sources of fat rather than raise its total, as milk, eggs, and nuts rise while meat and oil fall, leaving overall fat intake roughly unchanged (Section 5.1); the higher fat share under heat therefore reflects falling carbohydrate and protein intake rather than rising fat intake. Both hot and cold stress, therefore, shift individuals toward high-fat diets, with implications for future food demand and nutrition under climate change.

### 5.3 Potential mechanisms

The baseline coefficients in Section 5.1 summarize a reduced-form dietary response to short-run temperature exposure. They combine several channels that the design does not separately identify, such as physiological appetite regulation and changes in time use and activity. This subsection asks which of these channels the evidence can and cannot rule in. The goal is not to attribute the entire response to one mechanism, but to document which components are consistent with each channel and which are not.

The pattern of macronutrient responses is consistent with the two physiological forces summarized in the Conceptual framework. Heat activates pathways that suppress appetite and reduce total energy intake (Institute of Medicine, 1993), and carbohydrates and protein carry higher diet-induced thermogenesis during digestion than fat (Westerterp, 2004). Thus, selective suppression of carbohydrates and protein relative to fat raises the fat share of energy even when total consumption falls. Cold activates sympathetic thermogenesis, including shivering and brown-adipose-tissue (BAT) thermogenesis (Virtanen et al., 2009), which raises caloric requirements and favors energy-dense foods at 9 kcal per gram. Our decomposition shows carbohydrates and protein falling more than fat under heat and fat rising more than other macronutrients under cold. The physiology does not mechanically deliver these results, because prices, preferences, and adaptation capacity also shape dietary choice; we therefore describe the evidence as consistent with, rather than driven primarily by, physiological appetite regulation.

Two complementary tests help narrow the set of viable explanations. First, heat and cold may influence dietary behavior by altering individuals’ time allocation or physical activity. High or low temperatures could shorten labor supply or reduce leisure activities such as outdoor exercise (Graff Zivin and Neidell, 2014; Harris, 2026), indirectly affecting energy and macronutrient requirements. We examine this channel in two steps. We first note that, since occupation type and long-run physical activity preferences are time-invariant characteristics for most respondents, they are largely absorbed by the individual fixed effects already included in the baseline specification and thus provide limited additional identifying variation (for details about these variables, see Appendix B). Table A9 shows that the baseline temperature effects on dietary fat outcomes remain robust after controlling for work type and physical activity preferences, suggesting these channels are not the primary mediators.

Second, we directly examine whether temperature fluctuations affect working hours by regressing reported hours worked during the previous week on the corresponding week’s temperature exposure. As shown in Table A10, exposures to both heat and cold significantly reduce weekly working hours. This decline does not, however, resolve whether the response is physiological. If lower activity reduces energy expenditure, a person who eats less carbohydrate and protein may be matching intake to a lower requirement rather than undergoing a pathological shift, and the physiological and energy-balance accounts predict the same fall in carbohydrate and protein and the same rise in the fat share. Our design cannot separate them, and because both heat and cold reduce hours worked, expenditure plausibly moves in the same direction as intake, so we cannot sign the net effect on energy balance. The cold side is more informative here: hours worked also fall under cold, which lowers activity-related expenditure, yet total intake rises, so an activity-based energy-balance account alone cannot generate the cold-side response; the rise is more naturally attributed to thermoregulatory expenditure. We therefore treat consistency with appetite regulation as the strongest claim the evidence supports. Splitting the sample by occupational physical intensity points the same way. Responses are larger for physical laborers than for white-collar workers under cold, while under heat the residual, largely non-working group responds at least as strongly, so the heat response does not appear to require workplace exposure (Appendix F and Table A16). Ruling out an increase in physical labor narrows the behavioral explanations without establishing a physiological channel, and other demand-side channels such as food availability, prices, and cooking effort remain outside what our design can separate.

### 5.4 Heterogeneity and adaptation

In this section, we examine how these effects vary across population subgroups and what factors may attenuate them. Figure 2 (and Table A11) presents heterogeneity results across four dimensions.

**Figure 2.**
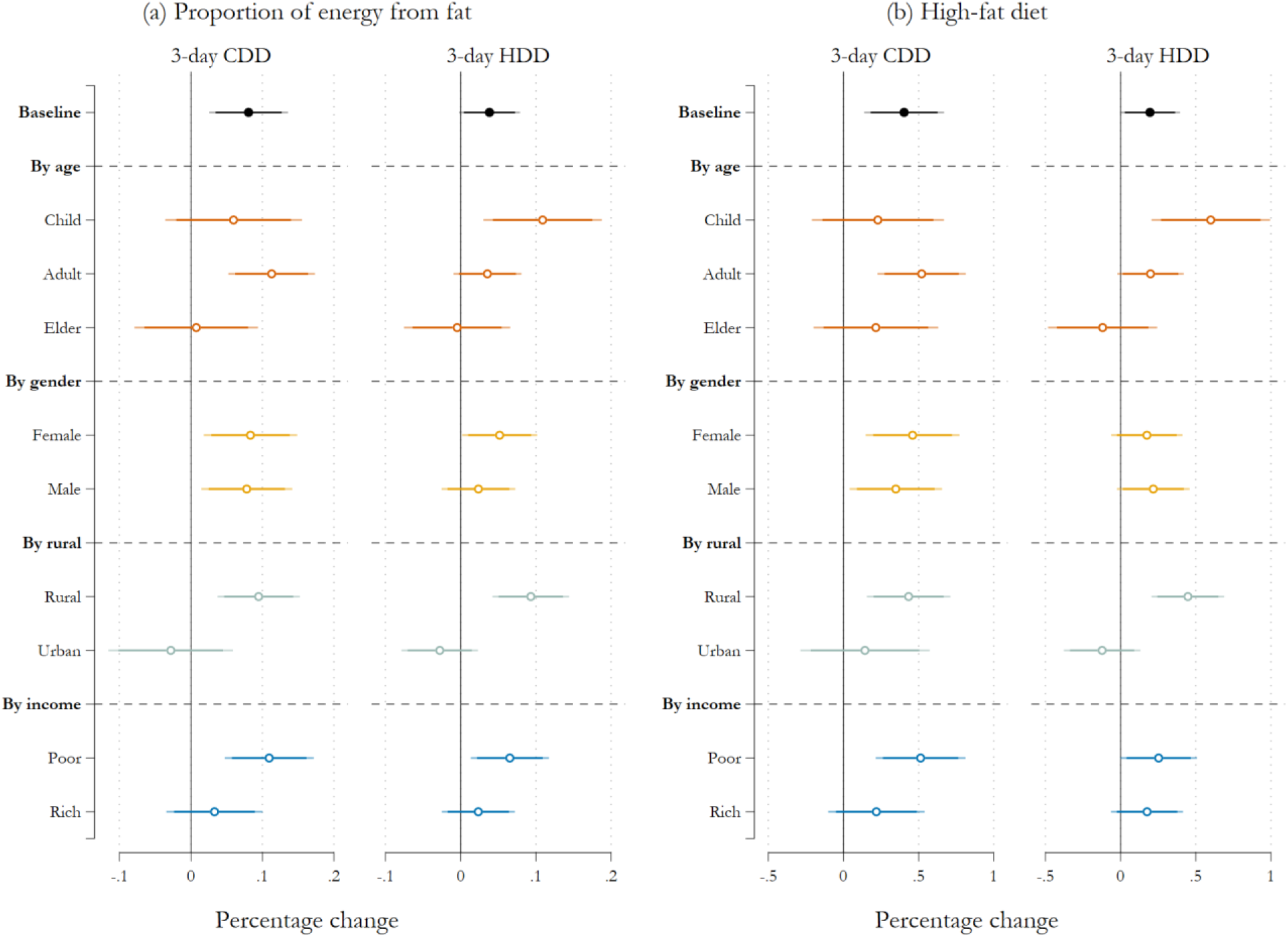
Heterogeneous effects of temperature on the proportion of energy from fat and high-fat diets. *Notes:* These figures show the heterogeneity of the effect of off-comfort temperatures on the proportion of energy from fat and the probability of having a high-fat diet. The outcome variables are the original levels. The regression model includes Individual fixed effects, year-month fixed effects, province-year fixed effects, and province-month fixed effects. The regression results are reported in Table A11. The circles represent the regression coefficients, and the dark (light) lines represent 90% (95%) confidence intervals. For ease of interpretation, we multiply the regression coefficients and CIs by 100 so that the result can be interpreted as a change in percentage. Standard errors are clustered at the individual level.

First, we focus on the heterogeneity of age and gender. Off-comfort temperatures affect older adults less than other age groups, possibly because age-related reductions in appetite buffer their physiological responses to thermal stress. Children, by contrast, appear more sensitive to cold than to heat. While dietary fat is an essential macronutrient during childhood development, the cold-induced shift toward high-fat diets documented here reflects a compositional change driven by thermal stress rather than deliberate nutritional planning. Given that early-life macronutrient intake shapes long-run health trajectories and human capital accumulation (Maluccio et al., 2009; Almond et al., 2015; Chong et al., 2016; Hjort et al., 2017; Coffey et al., 2018), repeated exposure to cold-induced dietary imbalances during childhood may reinforce unhealthy eating patterns with lasting consequences.

Second, rural populations are substantially more sensitive to temperature shocks than urban residents. Under both heat and cold, rural individuals experience significantly larger increases in the proportion of energy from fat and a higher likelihood of adopting a high-fat diet. This disparity reflects compounding structural disadvantages. Rural areas face inadequate food storage infrastructure, limited market access, and higher transportation costs (Liu et al., 2014), all of which constrain dietary flexibility. As shown in Table A12, access to cooling and heating appliances is also considerably lower among rural households, leaving them with fewer resources to buffer the physiological effects of thermal stress.

Third, dividing the sample at the median household income for each survey year, we find that individuals from lower-income households are notably more vulnerable to off-comfort temperatures. Both heat and cold drive larger increases in the fat share of energy and the probability of a high-fat diet among poorer individuals, while the effects among higher-income groups are smaller and often statistically insignificant. Wealthier households benefit from broader economic resources, including better access to cooling and heating technologies (Table A12).

The heterogeneity results above suggest that wealthier and urban households are better insulated from temperature-driven dietary shifts, and differences in appliance ownership are one plausible channel through which these advantages operate. To examine this channel more directly, we estimate equation (3) by interacting temperature with household ownership of fans, air conditioners, heating systems, and refrigerators (see Appendix G).^12^

Results are shown in Figure 3 and Table A13. Consistent with the heterogeneity patterns above, fan ownership significantly attenuates the heat-induced increases in the proportion of energy from fat and the probability of a high-fat diet. Air conditioner ownership shows no significant interaction with the dietary fat share or high-fat diet outcomes, which may reflect the low penetration of air conditioners in our sample (average ownership rate of 13.7%), where fans remain the dominant cooling device. On the cold side, heating system ownership partially reduces the cold-induced rise in the fat share, though the estimate is only marginally significant. Refrigerators, which mitigate temperature effects through food storage and availability rather than thermal comfort, show no significant interaction with either CDD or HDD on dietary fat outcomes. Notably, our data capture appliance ownership rather than actual usage. If owned appliances are not actively used, our estimates likely understate the mitigating effect of adaptation among households that actually use them.^13^

**Figure 3.**
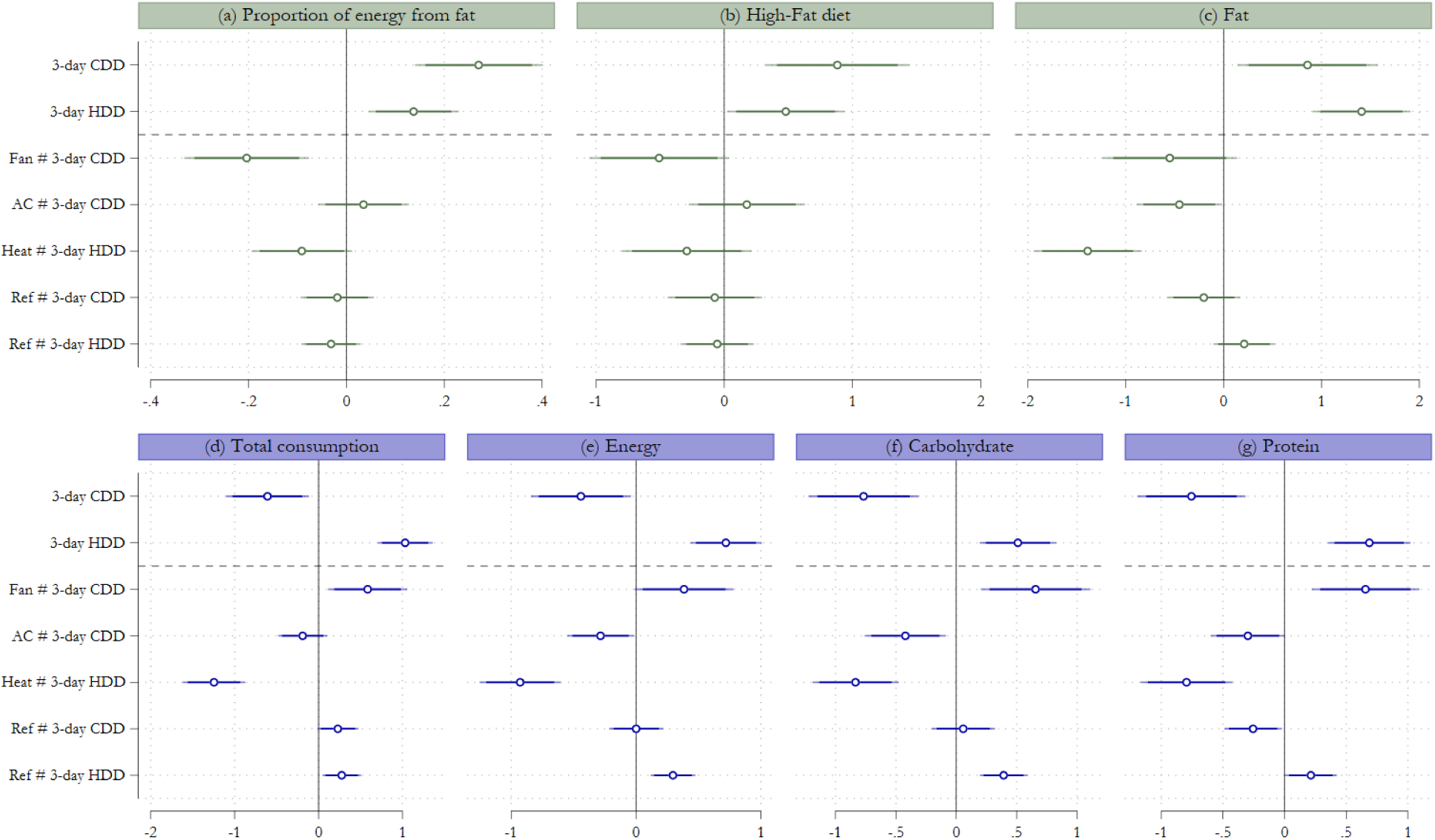
Adaptation effects of fans, air conditioners, heating systems, and refrigerators. *Notes:* These figures demonstrate the adaptation effects of home appliances on the proportion of energy from fat, high-fat diet index, 3-day food consumption, and macronutrient intake. The outcome variables are the logarithmic form of the value of food consumption or macronutrient intake and the original levels of the proportion of energy from fat and high-fat diets. As shown in equation (3), the regression model includes Individual fixed effects, year-month fixed effects, province-year fixed effects, and province-month fixed effects. The regression results are documented in Table A13. The circles represent the regression coefficients, and the dark (light) lines represent 90% (95%) confidence intervals. For ease of interpretation, we multiply the regression coefficients and CIs by 100 so that the result can be interpreted as a change in percentage. Standard errors are clustered at the individual level.

Taken together, these findings suggest that the dietary consequences of temperature stress are not uniformly borne. Structurally disadvantaged groups, including rural residents and lower-income households, face the greatest exposure and the fewest resources to adapt. While household appliances represent one mechanism through which better-off households buffer dietary risks, addressing the broader socioeconomic conditions that generate these vulnerabilities is equally important from a policy standpoint.

## 6. Conclusion

Climate change is increasing the frequency and intensity of off-comfort temperatures globally. Using detailed longitudinal data on individual-level dietary intake combined with high-resolution weather records, this paper documents that short-term temperature deviations from comfort systematically shift dietary composition toward higher fat share. Both hot and cold temperatures raise the dietary share of energy from fat and increase the likelihood of adopting a high-fat diet. While we cannot test whether short-run absolute fat increases are fully offset by thermogenic expenditure, the clinical literature associates a higher fat share with metabolic risks (e.g., impaired insulin sensitivity and adverse lipid profiles) independent of total calorie changes.

Two implications follow for how economists account for the welfare cost of climate change. First, nutrition belongs alongside mortality, morbidity, labor supply, and agricultural output as a margin through which temperature affects health capital. Calorie-based adequacy measures can miss a deterioration in diet quality when the mix of macronutrients moves toward fat, and the same movement can occur whether total calories fall under heat or rise under cold. Second, the distributional signature of the dietary response implies that the welfare cost of climate change is not equally shared. Rural residents and lower-income households absorb larger compositional shifts, and appliance ownership correlates with smaller shifts, suggesting that these patterns reflect adaptive capacity rather than the causal effect of any single technology. The importance of this capacity is further underscored by our end-of-century projections (Appendix H). Although inherently subject to strong structural assumptions, these projections suggest a plausible scenario in which the dietary consequences of short-run temperature deviations intensify substantially in the absence of targeted adaptation efforts. Quantitatively, under RCP8.5 the projected end-of-century increase averages about 0.2 to 0.3 percentage points in the fat share of energy and about 1 to 1.5 percentage points in the probability of a high-fat diet, equivalent to roughly half a year of the historical secular trend in the fat share, with substantially larger shifts in the hot southern regions.

Several directions remain for future work. First, linking short-run dietary responses to panel measures of body composition and clinical outcomes would test whether the compositional shifts documented here translate into cardiometabolic risk, a step our data cannot complete. Second, extending the design to food-price microdata would separate demand-side adjustment from high-frequency supply-side channels more sharply than growing-season controls can. Finally, our estimates capture short-term behavioral responses. Whether these transient adjustments accumulate into persistent long-run dietary shifts under sustained warming remains an open question, introducing uncertainty into our projection exercises. Because our identification deliberately isolates short-run deviations and holds constant the slower margins through which households adapt, the estimated responses are best read as immediate reactions to weather shocks rather than direct forecasts of how several decades of warming will reshape diets. Overall, our results point to individual diet as a margin on which climate and inequality interact, and they locate this margin inside the broader economic analysis of climate and health capital rather than outside it.

## Data Availability

The dataset used in this paper contains confidential information from the China Health and Nutrition Survey (CHNS) that can be used to identify respondents' locations. Therefore, the authors cannot share the full dataset. However, readers may download part of the dataset from the CHNS official website.

https://www.cpc.unc.edu/projects/china/

## Appendix A: Conceptual framework

An individual chooses daily intakes of carbohydrate *c*, protein *p*, and fat *f* (in grams) to maximize a consumption value

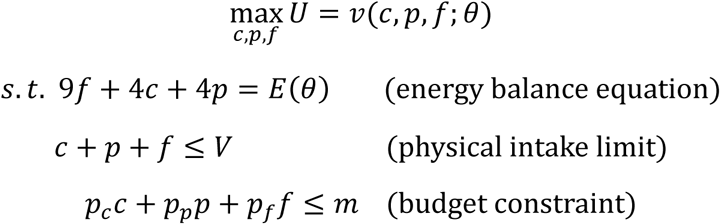

where *θ* is ambient temperature, *V* is the maximum feasible intake, *p_c_* , *p_p_* , and *p_f_* are the corresponding prices, and *m* is the food budget. The central object is the fat share of energy

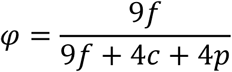

with fat contributing 9 kcal per gram and carbohydrate and protein 4 kcal each.

We treat *v*(·; *θ*) as a single reduced-form consumption value that combines the physiological appetite signal and the hedonic taste for each macronutrient. Let *v_c_* ≡ *∂v*/*∂c*, *v_p_* ≡ *∂v*/*∂p*, and *v_f_* ≡ *∂v*/*∂f* denote the marginal consumption values of carbohydrate, protein, and fat. Our data cannot separate a physiological drive from a psychological one, and the biomedical literature links thermal stress to both in the same direction, so representing them as one temperature-dependent preference shifter is a modeling choice rather than an identifying assumption.

Temperature enters through two channels. (i) A thermogenic-comfort margin in *v*. Digesting food releases heat. Diet-induced thermogenesis (DIT) dissipates 20-30% of protein energy and 5-10% of carbohydrate energy, versus only 0-3% for fat (Westerterp, 2004). This metabolic heat is pleasant under cold but aversive under heat, so *∂v_c_*/*∂θ* < 0 and *∂v_p_*/*∂θ* < 0 relative to *∂v_f_*/*∂θ*, and high-DIT carbohydrate and protein become comparatively less attractive as temperatures rise. (ii) The energy requirement *E*(*θ*) . Cold activates sympathetic thermogenesis, shivering and brown-adipose-tissue oxidation, raising the body’s energy need (Virtanen et al., 2009), while heat suppresses appetite and lowers it.

These primitives generate two opposing margins that both raise the fat share.

**Heat (*θ* high).** Margin (i) dominates. The thermogenic load of carbohydrate and protein becomes aversive, inducing substitution toward fat, while a lower *E*(*θ*) reduces total intake. Carbohydrate and protein fall more sharply than fat, whose absolute intake remains roughly unchanged, so *φ* rises. This is a composition (denominator) effect.

**Cold (*θ* low).** Margin (ii) dominates. The elevated requirement *E*(*θ*) must be met under budget and physical intake limits; because fat is the most energy-dense macronutrient, the requirement is met most efficiently by raising fat intake. Fat rises more than carbohydrate and protein, so *φ* rises. This is an absolute-intake (numerator) effect.

The framework thus predicts a rising fat share at both hot and cold temperature, but through distinct margins. Heat operates as a composition effect and cold as an absolute-intake effect, and this asymmetry is what we test in Section 5.

## Appendix B: Data cleaning

### B1. Food consumption

Detailed food consumption information is collected using the 24-hour recalls on 3 consecutive days, with each food item assigned a corresponding food code from the Chinese Food Composition Table (FCT). Household food consumption was measured daily by weighing all available foods at the start and end of each survey day, alongside detailed recording of purchases, home production, and food waste. Individual dietary intake data were gathered each day through interviews, capturing both at-home and away-from-home food consumption.^14^ The household and individual dietary intakes were cross-validated to ensure data quality, with trained nutritionists conducting all interviews (China Health and Nutrition Survey Data Collection, 2020).

To derive the average daily food consumption, we aggregate individual-level food consumption over the 3-day interview period and divide it by the total person-days for each type of food. Any observations with missing food codes are excluded from the analysis. For individuals with multiple records on the same day, we consider the highest number of person-days as the count. Only 1,512 of 3,040,836 observations have this problem. In the raw data, 2,075,211 observations have the records of person-days, and approximately 95.7% of them are recorded as 1. Missing values occur throughout all years, so we substitute the incorrect person-days with the value of 1. It is worth noting that while most individuals have completed 3-day records, some only have 1-day or 2-day records. We opt to drop these observations for two reasons. First, food consumption data from individuals with fewer than three days of records raises reliability concerns. Second, 1-day or 2-day records are more prone to measurement errors compared to 3-day records, which could potentially bias the estimation. We drop 27,792 of 3,040,836 observations in this step.

During the survey periods, three versions of the FCT were available, requiring the alignment of food codes with specific items. We classify all items into 12 distinct food categories, primarily aligning them with the current version of the FCT. These 12 categories encompass cereals, tubers, dried legumes, vegetables, fungi, fruits, nuts, meats, poultry, dairy products, eggs, and seafood. To ensure data accuracy, we exclude observations with extreme values. Specifically, observations are omitted if the z-score for any food category exceeds 10. Furthermore, specific categories such as infant foods, ethnic cuisine, fast food, and beverages are disregarded due to the varying classifications and definitions across different versions of the FCT. For each person, we adjust their 3-day intake based on the person-days. We sum up all the 12 types of food consumed by the individual as one variable, i.e., total food consumption. Since CHNS only provides the total consumption amount of each household for the condiments, we average the total amount for everyone who eats at home.

We drop the data from the first and final surveys in our analysis for three reasons. First, the sample size in 1989 was relatively small compared with other waves. Only people aged 1-6 and 20-45 were included in the food consumption survey. Second, the first survey wave lasted from 1989 to 1990. Finally, the official CHNS website does not provide information on food consumption and macronutrient intake for 2015. Before using the data in empirical analysis, we exclude observations in which the key dependent variables (i.e., total food consumption, carbohydrate, protein, and fat) fall outside the 0.5th to 99.5th percentile range.

### B2. BMI

When handling BMI data, we carefully review the original entries and exclude observations suspected of containing erroneous height or weight values, as well as those of juvenile individuals. Furthermore, observations with BMI values falling outside the 0.5th to 99.5th percentile range are removed from the analysis.

### B3. Working and exercising

For working and exercising variables, most survey questions refer to annual averages, presenting challenges in accurately aligning these responses with short-term temperature fluctuations.

First, to capture exercise behaviors, we construct a variable named “sport time” by aggregating respondents’ daily time spent engaging in various physical activities. These activities include martial arts, gymnastics, dancing, acrobatics, running (track and field), swimming, soccer, basketball, tennis, badminton, volleyball, and other forms of exercise such as ping pong and Tai Chi. Additionally, we create three dummy variables, “walking”, “exercise”, and “bodybuilding”, to represent individual preferences for these specific physical activities. Given the lack of precise time frames associated with these variables, we include them solely as control variables in our mechanism analysis.

Second, concerning work-related data, we address the timing mismatch by using responses to the survey question: “During the past week, for how many hours did you work?” This includes working hours from both primary and secondary occupations. We then match each individual’s reported working hours with their corresponding temperature exposure during the preceding week, based on precise interview dates. To minimize measurement errors, evident outliers, such as weekly working hours exceeding the 99.5th percentile (e.g., 168 hours), were excluded from our analysis.

## Appendix C: Balance test of interview timing

A natural concern is whether the assignment of interviews to high-CDD or high-HDD days is quasi-random conditional on the fixed effects, given that fieldwork logistics might prioritize more accessible households under extreme weather. We address this in two steps. We first test whether predetermined characteristics predict temperature exposure, and we then show that the main estimates are unchanged when we control for the characteristics on which any imbalance appears.

### C1. Balance of predetermined characteristics

Table A14 regresses four predetermined characteristics (urban status, sex, age, and an indicator for above-wave-median household income) on the 3-day CDD and HDD in four separate regressions, together with the baseline weather controls. Panel A includes year-month, province-year, and province-month fixed effects; Panel B adds individual fixed effects, matching our baseline specification. The joint F-test rows come from a separate regression that reverses this direction: within each panel, we regress the 3-day CDD (or HDD) on all the characteristics in that panel jointly, with the same fixed effects, and test whether the characteristics are jointly zero. The column regressions therefore ask whether temperature predicts each characteristic one at a time, whereas the joint test asks whether the characteristics together predict the temperature of the interview window.

Without individual fixed effects (Panel A), interview timing is not perfectly balanced. Hotter survey windows draw somewhat more rural, younger, and lower-income respondents, and the four characteristics jointly predict both CDD and HDD (joint F-tests in Table A14). Our baseline, however, includes individual fixed effects, which absorb all time-invariant characteristics (rural status and sex). Panel B repeats the test with individual fixed effects. Sex and rural status drop out by construction, and the cold-side coefficient on age remains statistically significant, although it is tiny (0.0014 years per degree-day). The above-median income indicator retains a small residual correlation with temperature, and age and income together still predict temperature in the joint test. The income indicator also changes sign between the two panels, and the two estimates describe different comparisons. Panel A compares different respondents, and those interviewed in hotter windows tend to come from lower-income households. Panel B compares the same respondent across survey waves, and a given respondent’s hotter interview windows tend to fall in the waves in which the household is above the median. The two need not agree, but the within-respondent pattern runs opposite to the concern that fieldwork under extreme weather favors easier-to-reach, better-off households. We cannot determine its source from our data. One possibility is that the timing of fieldwork within a county drifted across waves in a way that happens to coincide with households’ income trajectories, but this is a conjecture rather than a finding. Whatever the source, because income is correlated with dietary composition, any correlation between income and interview-window temperature could bias the temperature coefficients. If higher income goes with a higher fat share, the bias would be downward under the between-household pattern and upward under the within-respondent pattern that applies to our baseline. We therefore control for income directly in Table A15 and the estimates are essentially unchanged.

### C2. Baseline estimates with additional controls

Table A15 re-estimates the baseline effects on the proportion of energy from fat and the probability of a high-fat diet, progressively adding controls for per capita household income and age, with all columns estimated on the common sample so that they are directly comparable. Adding the controls leaves the estimates essentially unchanged. The CDD coefficients barely move (0.0716 to 0.0714 in Panel A and 0.3842 to 0.3817 in Panel B), and the HDD coefficients become slightly larger (0.0376 to 0.0379 in Panel A and 0.1881 to 0.1922 in Panel B). Statistical significance is unchanged throughout. The residual correlations documented in Table A14 are therefore detectable in a sample of this size but immaterial for the estimated temperature effects, since controlling for the characteristics directly does not move them. The cold side is particularly informative. Colder survey windows draw slightly lower-income respondents, which could in principle inflate the HDD estimate; the income control addresses exactly this channel, and adding it slightly increases rather than decreases the HDD coefficient, indicating that selection is not generating the cold-side effect.

## Appendix D: Correlation of BMI with energy intake and dietary structure

We use the OLS approach to reveal the correlation of BMI with energy intake and dietary structure.

Since BMI measurement in the CHNS could fall on any day within the 3-day dietary recall window, regressing it on contemporaneous dietary records introduces a timing mismatch. Moreover, short-term food intake records are difficult to link logically to BMI as a stock measure of nutritional status. We therefore compute each adult individual’s average fat intake and fat share across all survey waves as a proxy for their long-run dietary equilibrium, and use BMI measured at the last observed wave as the outcome variable in a cross-sectional regression. The model is as follows:

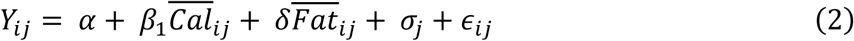

where *Y_ij_* represents the BMI of adult individual *i* (age ≥ 18) in community *j*, measured at their last observed survey wave. *Cal_ij_* is individual *i*’s average total calorie intake across all survey waves. *Fat_ij_* represents the fat-related variables, which include the real amount of fat intake, the proportion of energy from fat, or the probability of having a high-fat diet, each averaged across waves. *σ_j_* denotes community fixed effects, which absorb differences across communities. According to Chinese criteria, we further define an overweight indicator equal to one if an adult’s BMI is at or above 24, to give the results a clearer health interpretation. We further control for the averages of working hours and physical activity variables, which may be related to BMI and overweight across all survey waves, and the results remain consistent with the specification without these controls.

## Appendix E: Robustness checks

To verify the robustness of our findings, we conduct a series of tests. First, we replace the baseline fixed effects with individual fixed effects and province-year-month fixed effects, which impose a more stringent set of controls.

Second, we address the concern that survey-period temperatures may be correlated with growing-season temperatures within the same year, which could confound our demand-side estimates through agricultural supply-side channels. For instance, if an unusually hot summer reduces local crop yields and thereby alters food availability and prices during the survey period, our estimates may partly capture supply-side effects rather than the direct demand-side response to thermal stress. To isolate the demand-side effect, following Chen and Gong (2021), we augment the baseline specification with county-year GDD (10-33°C) and KDD (>33°C) accumulated during the main crop growing season, where the growing-season window is defined separately for each province based on its dominant staple crops and local agricultural calendar. This county-year measure captures local yield-relevant temperature shocks that remain after conditioning on province-year fixed effects, which only absorb province-level average conditions. We do not include these controls in the baseline specification because the province-specific growing-season windows are defined at the provincial level, while temperature itself varies at the county level, introducing a degree of measurement error that could add noise to the estimates; the province-year fixed effects already absorb the bulk of supply-side variation.

Third, we address the concern raised by Jones et al. (2026) that nonlinear temperature specifications may generate spurious effects when warming-induced trends in temperature exposure correlate with place-specific trends in outcome variables. For each county-month cell, we estimate a linear time trend in monthly mean temperature using daily weather data from 1981 to 2011 (thirty years in total), extending the sample back to 1981 to improve the precision of trend estimation, and shrinking the estimated slopes toward the cross-county mean using empirical Bayes techniques following Jones et al. (2026). To accurately capture the nonlinear effect of warming on degree days, we project the historical daily temperature distributions onto these estimated county-month warming trends. We then calculate the expected daily CDD and HDD based on these projected distributions and aggregate them to match the exact three-day survey window of each respondent. Including these window-specific expected measures as additional controls absorbs the mechanical variation driven by differential long-term climate trends, leaving the baseline three-day CDD and HDD identified strictly by idiosyncratic short-run weather shocks.

Fourth, we add day-of-week fixed effects to absorb systematic differences in diet between weekdays and weekends. Fifth, we add an indicator for major festival periods, equal to one within three days of the Spring Festival, the Dragon Boat Festival, or the Mid-Autumn Festival, or during the National Day holiday, to control for holiday-related dietary variation. Sixth, we drop all meteorological control variables other than temperature. All these regression results are reported in Table A6.

Seventh, we test alternative thresholds for CDD and HDD, including 4–26°C, 3–27°C, and 2–28°C. We also compute 3-day cumulative CDD and HDD using daily maximum and minimum temperatures. Because the distribution of daily maximum (minimum) temperatures is significantly shifted to the right (left) relative to the average temperature (as shown in Figure A1(c)), the cutoff for maximum temperature is adjusted to 30°C and for minimum temperature to 0°C. These results, shown in Table A7, are consistent with our baseline estimates.

## Appendix F: Temperature effects by occupational physical intensity

Physically demanding occupations expose workers more directly to ambient temperature than sedentary ones, so dietary responses should be larger for physical laborers if workplace exposure matters. Because the working-hours response in Table A10 is estimated on employed respondents, most of whom are physical laborers, the energy-balance channel also applies most directly to this group; we therefore read this contrast as an exposure heterogeneity test, which does not by itself separate physiological regulation from the energy-balance channel, since both predict larger responses for the more exposed group. We split the sample by primary occupation into physical laborers (farmers and skilled and unskilled manual workers), white-collar workers (professionals, administrators, and office staff), and a residual group of all remaining respondents, namely those in other occupations and those not reporting an occupation (including children), so that the full sample is retained. We then interact the 3-day CDD and HDD with these three groups.

Table A16 reports the temperature coefficients by occupational group. The temperature slopes for physical laborers are significant under both heat and cold in both columns, whereas the corresponding coefficients for white-collar workers are uniformly insignificant. The point estimates for physical laborers exceed the white-collar ones under cold in both columns and under heat for the high-fat diet indicator, while the heat slopes for the fat share are essentially the same in the two groups. A formal Wald test of equality between the physical-labor and white-collar slopes rejects at the ten percent level only for the fat share under cold (p=0.079) and does not reject in the other three comparisons, so the contrast is in the expected direction, and clearest under cold, but not statistically decisive.

The heat response is present both for physical laborers and for the residual group, which mainly consists of respondents without a reported occupation, so it does not appear to require occupational exposure; this is in line with the appetite interpretation of the hot-side channel in Section 5.3. Two qualifications apply. The white-collar group is small (about eight percent of the sample), so its insignificant coefficients are also imprecisely estimated, and the Wald tests are at best marginally significant. Moreover, occupation is not randomly assigned. Physical laborers differ from white-collar workers along many dimensions that are themselves associated with the size of the dietary response (see Figure 2), most importantly wealth, and rural residence. We therefore read this table as suggestive.

## Appendix G: Adaptation

The adoption of these home appliances may be endogenous. Extreme temperatures may affect the work time (Somanathan et al., 2021; Heyes and Saberian, 2022) and income (Carleton, 2017) of individuals, which determines the household purchasing power. Therefore, we use information on the respondents’ initial per capita household income by year fixed effects to control for the impact of wealth. All the income variables are deflated by the management of CHNS, so they are comparable even though different households may enter the survey in different waves.

Since the CHNS questionnaire does not document the adoption of district heating systems in households, we instead rely on whether the respondent’s county provided collective heating services. Specifically, households are defined as having heating systems if their county offered collective heating services (i.e., in the north of the Qinling-Huaihe line) during the survey period from November to February. The regression model employed in this section is as follows:

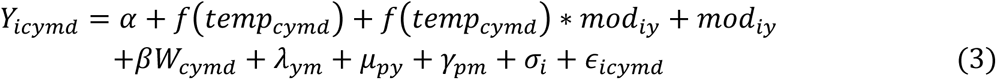

where, *mod_iy_* represents a vector of the technologies of individual *i* in year *y*, including whether there are fans, air conditioners, heating systems, and refrigerators in the household. If the answer is yes, the dummy variable *mod_iy_* is 1; otherwise, 0. Because the fans, air conditioners, and heating systems are primarily used during the summer and winter seasons, respectively, we interact these indicators with CDD (for air conditioners and fans) and HDD (for heating systems). Air conditioners may be used for both heating and cooling. However, given the early period of our study and the overall low rate of air conditioning ownership, we argue that people used air conditioning primarily for cooling during the study period. The remaining variables and symbols are the same as those in equation (1).

## Appendix H: Projection method and end-of-century projections

Climate projections predict that the global climate will experience more extremely hot days and fewer extremely cold days by the end of this century (see Figure A7(b)). In this context, we focus on two climate scenarios, RCP4.5 and RCP8.5: the former is a medium stabilization pathway assuming some global action to reduce emissions, while the latter is a high-concentration pathway assuming little effort to mitigate climate change (van Vuuren et al., 2011). Under these two scenarios, we project end-of-century temperature-induced changes in the proportion of energy from fat and the probability of adopting a high-fat diet, based on empirical relationships from equations (1) and (3).

We use the climate projection data provided by the NASA Earth Exchange Global Daily Downscaled Projections (NEX-GDDP) to predict the impacts of climate change on food consumption and macronutrient intake by the end of this century (2080-2099). The NEX-GDDP dataset includes downscaled projections for four Representative Concentration Pathways (RCPs) from 21 models conducted under the Coupled Model Intercomparison Project Phase 5, developed to support the Fifth IPCC report. We use RCP4.5 (an emissions stabilization scenario) and RCP8.5 (a scenario with intensive growth in fossil fuel emissions, namely “business-as-usual”) in line with the literature (Li et al., 2019; Carleton et al., 2022; Lai et al., 2022). The dataset contains daily maximum and minimum temperatures at a spatial resolution of 0.25 degrees. Our projection is based on the median projected temperature and climate from the 21 models and aggregates the original dataset to the city level daily temperature.

We predict the overall impact of temperature changes based on our baseline regression results (in Table A3) for each city *j*, using the model as follows:

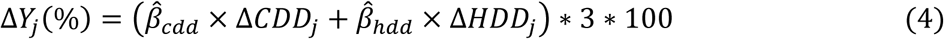

where Δ*Y_j_*(%) indicates the temperature change impacts on food consumption and macronutrient intake of individuals in city *j*. β^*_cdd_* and β^_ℎ*dd*_ represent the estimated coefficient for 3-day CDD and 3-day HDD in our baseline regression, respectively. Δ*CDD_j_* and Δ*HDD_j_* are the predicted change of yearly-average 1-day CDD and HDD in city *j*. Here we consider the changes between our survey waves (i.e., 1991, 1993, 1997, 2000, 2004, 2006, 2009, and 2011) and 2080-2099.

We also consider the effect of adaptation strategies. This projection scenario assumes that everyone simultaneously has a fan, refrigerator, and heating system in the household. Not all individuals may be equipped with all adaptation methods (e.g., residents in South China may have no heating systems). Thus, the result in this scenario is a lower bound of the actual effect of off-comfort temperatures because we may overestimate the effects of adaptation behavior at the aggregate level. The model is as follows:

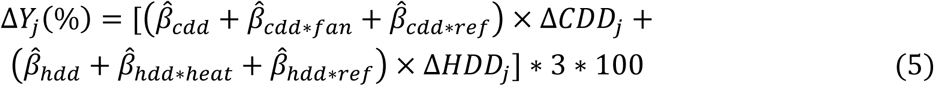

represent the coefficient of 3-day CDD by fans in Table A13. β^*_cdd_*_∗*ref*_ represents the coefficient of 3-day CDD by refrigerators, and β^_ℎ*dd*∗*ref*_ means the coefficient of interaction of 3-day HDD and refrigerators. β^_ℎ*dd*∗ℎ*eat*_ represents the coefficient of 3-day HDD by heating systems. These four coefficients capture the effects of adaptation strategies. In implementing this projection, we exclude the air-conditioner interaction, because its estimated coefficient is positive rather than protective for the two outcomes we project, the proportion of energy from fat and the high-fat diet indicator. Air conditioning shows no detectable mitigating effect against temperature-driven dietary shifts in our sample, though it remains in the estimating regression as a control.

These two equations correspond to two adaptation strategies: (1) the current level, where adaptation remains at the status quo, and (2) the complete adaptation. While economic growth will likely increase the adoption of fans, air conditioners, and other heating products, complete saturation remains implausible. Our projections thus establish upper (baseline) and lower (complete adaptation) bounds for climate-driven dietary impacts.

Before presenting the projection results, we emphasize that the results below should be interpreted with considerable caution. Our baseline specification identifies the effect of short-run (3-day) off-comfort temperature exposure on contemporaneous dietary composition, exploiting quasi-random within-individual variation. Translating these short-horizon estimates into end-of-century projections requires extrapolating along several dimensions simultaneously: from transitory shocks to sustained shifts in the temperature distribution, from the historical climate observed in our sample to climates that lie partly outside this support, and from the current socioeconomic environment to one that may differ materially in food markets, technology, and preferences. Each of these steps introduces assumptions that our identification strategy is not designed to validate.

The results shown in Figure A7(a) have three observations. First, individuals in more than half of the areas are likely to consume excess fat and face a higher risk of high-fat diets due to global warming, with these effects more pronounced under RCP8.5 than under RCP4.5. Second, while the average effects appear modest, the spatial distribution reveals considerable regional heterogeneity. Overall, southern China exhibits greater vulnerability, with individuals more prone to shifting toward high-fat diets. In the Northeast, the risk of high-fat diets declines, a pattern likely driven by the reduction in cold weather while temperatures remain in a relatively cool rather than hot range. Third, adaptation technologies substantially mitigate the dietary effects of temperature change, particularly for the probability of adopting a high-fat diet. Under complete adaptation, the projected increases in both the proportion of energy from fat and the probability of high-fat diets are reduced to approximately half those under the current-level scenario for both RCP4.5 and RCP8.5. This implies that adaptation strategies offset nearly half of the climate-induced dietary impact, underscoring the importance of expanding adaptive capacity for improving population dietary health.

## Appendix Figures

**Figure A1.**
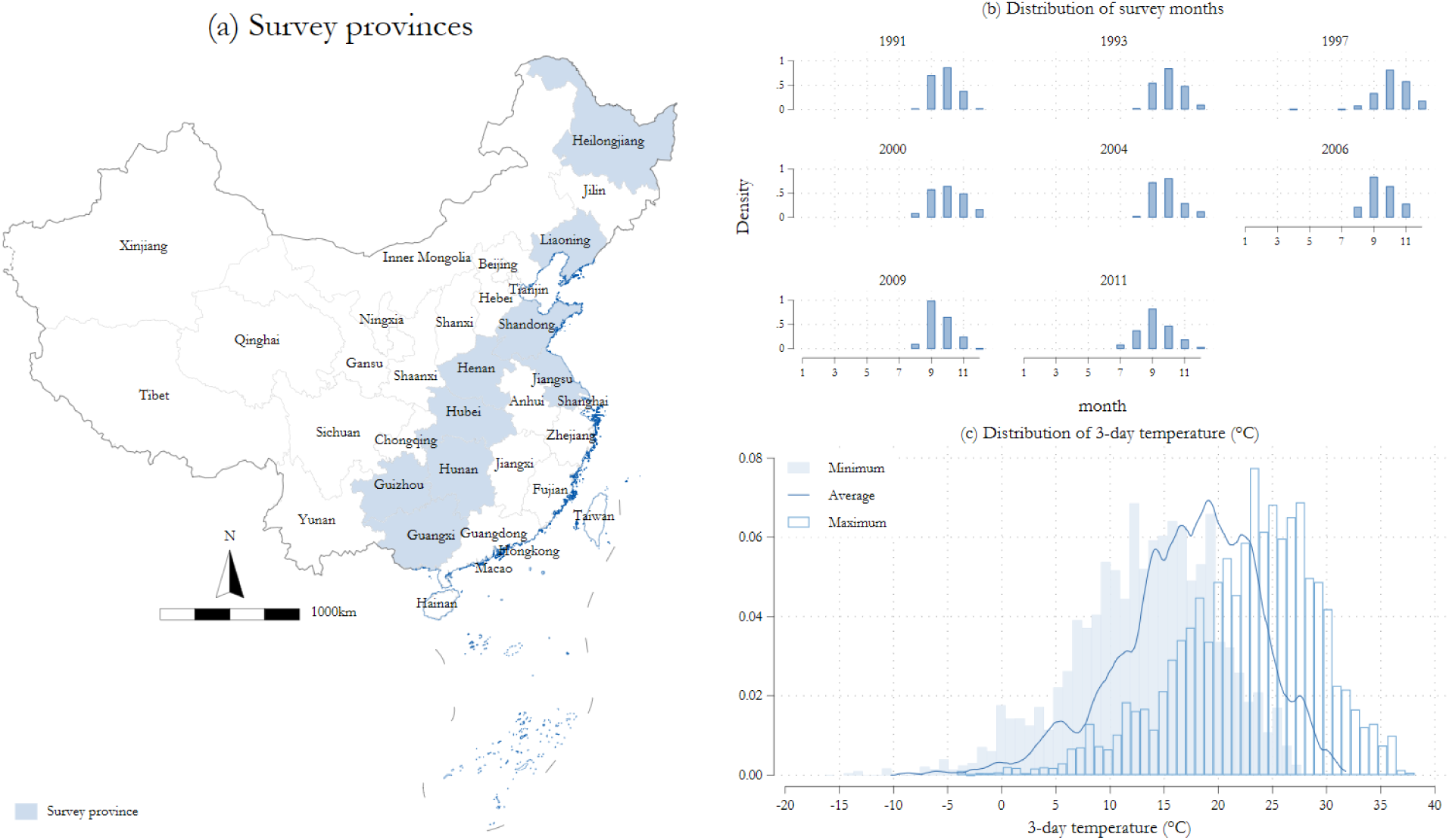
**Survey provinces, months, and the distribution of 3-day temperature.***Notes:* These figures show the survey provinces used in this paper (in (a)), the distribution of survey months in each year (in (b)), and the distribution of the 3-day temperature exposure by respondents (in (c)). The average temperature is concentrated between approximately 0°C and 30°C, with only a small fraction of observations outside this range, because the survey months mainly concentrated on September to November. The distribution patterns for maximum and minimum temperatures are similar to those of the average temperature, albeit with an overall shift in the distribution.

**Figure A2.**
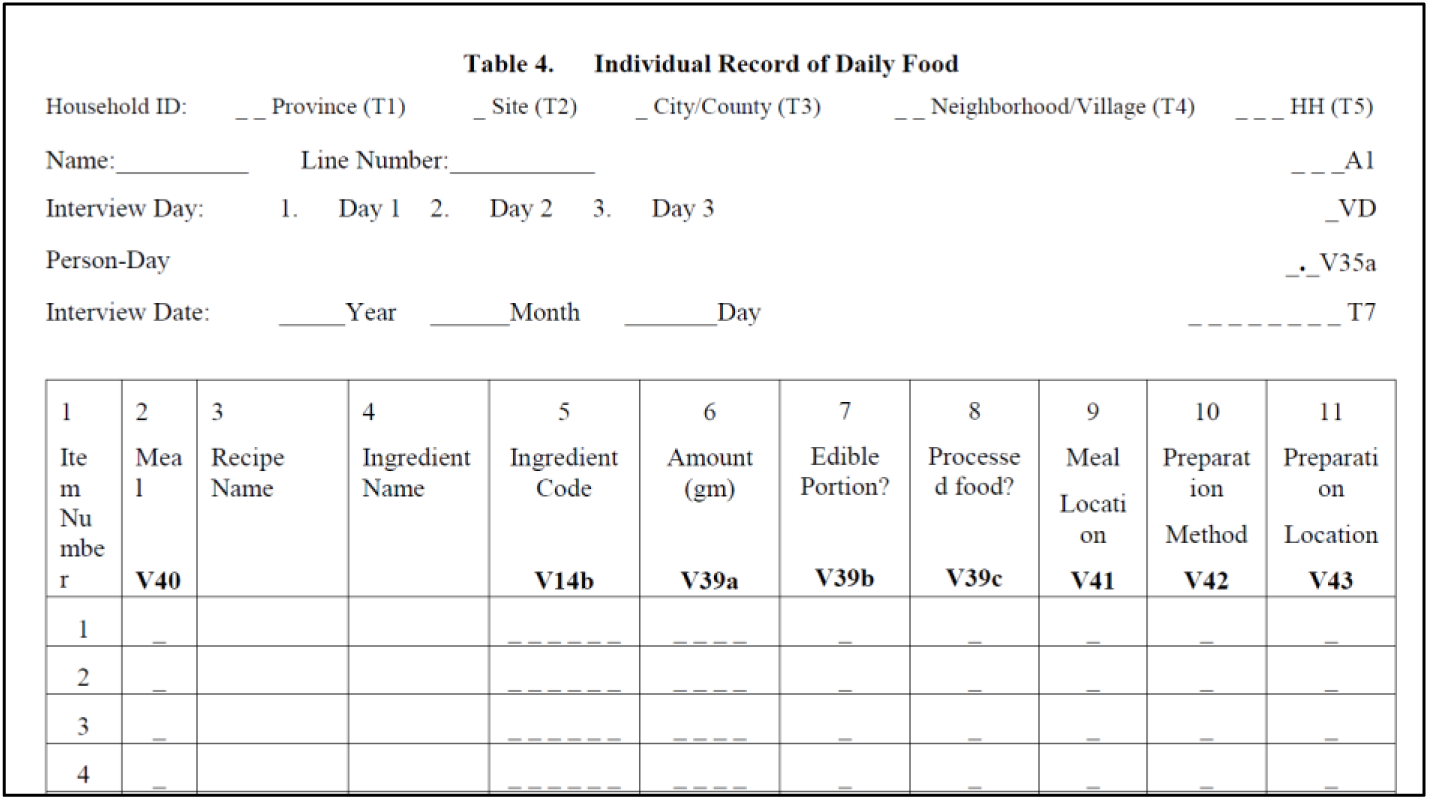
CHNS questionnaire: Food consumption (2011 version). *Notes:* This figure shows part of the CHNS questionnaire about food consumption.

**Figure A3.**
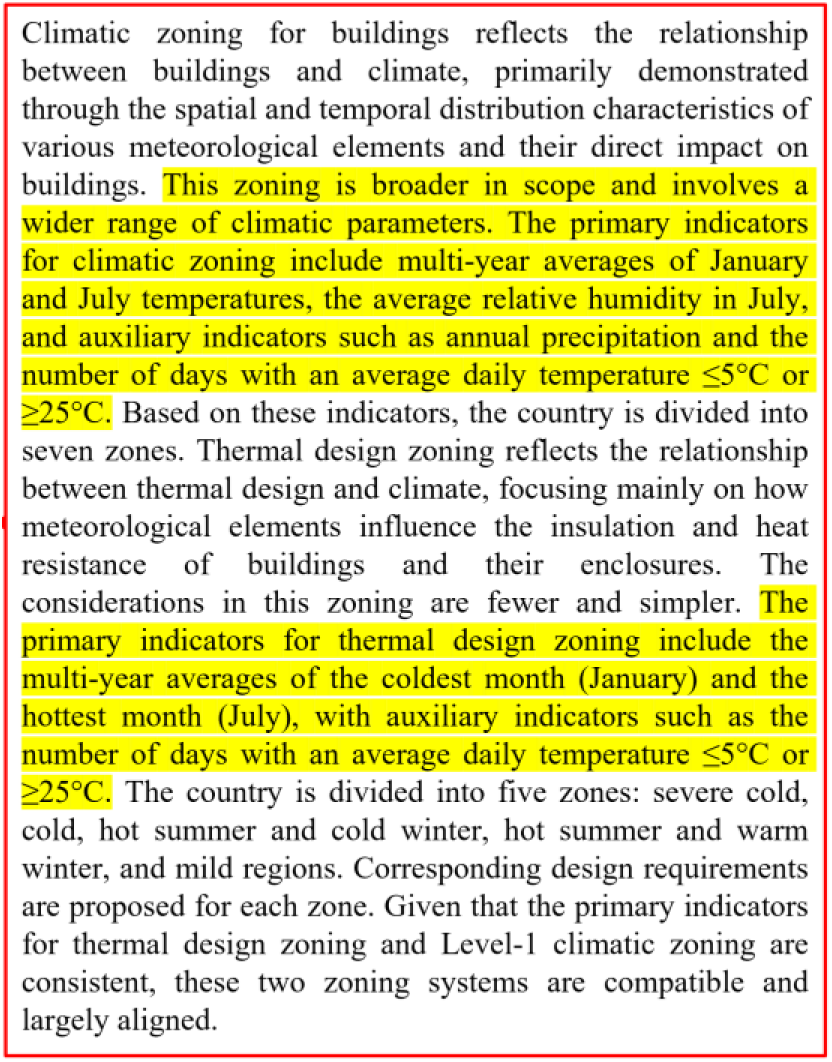
**Uniform standard for design of civil buildings (GB 50352-2019), page 71.***Notes:* This picture shows part of the *Uniform standard for design of civil buildings (GB 50352-2019)*. The main information in this picture is that “when zoning the climate region, we usually use the annual average temperature in January and July and the average relative humidity in July as the main indicators. The annual precipitation and the number of days with the average daily temperature of ≤5°C and ≥25°C are auxiliary indicators……When considering the building’s thermal design, the average temperature of the coldest month (that is, January) and hottest month (that is, July) is used as the main index of the zoning, and the number of days with average daily temperature ≤5°C and ≥25°C are auxiliary indicators.”

**Figure A4.**
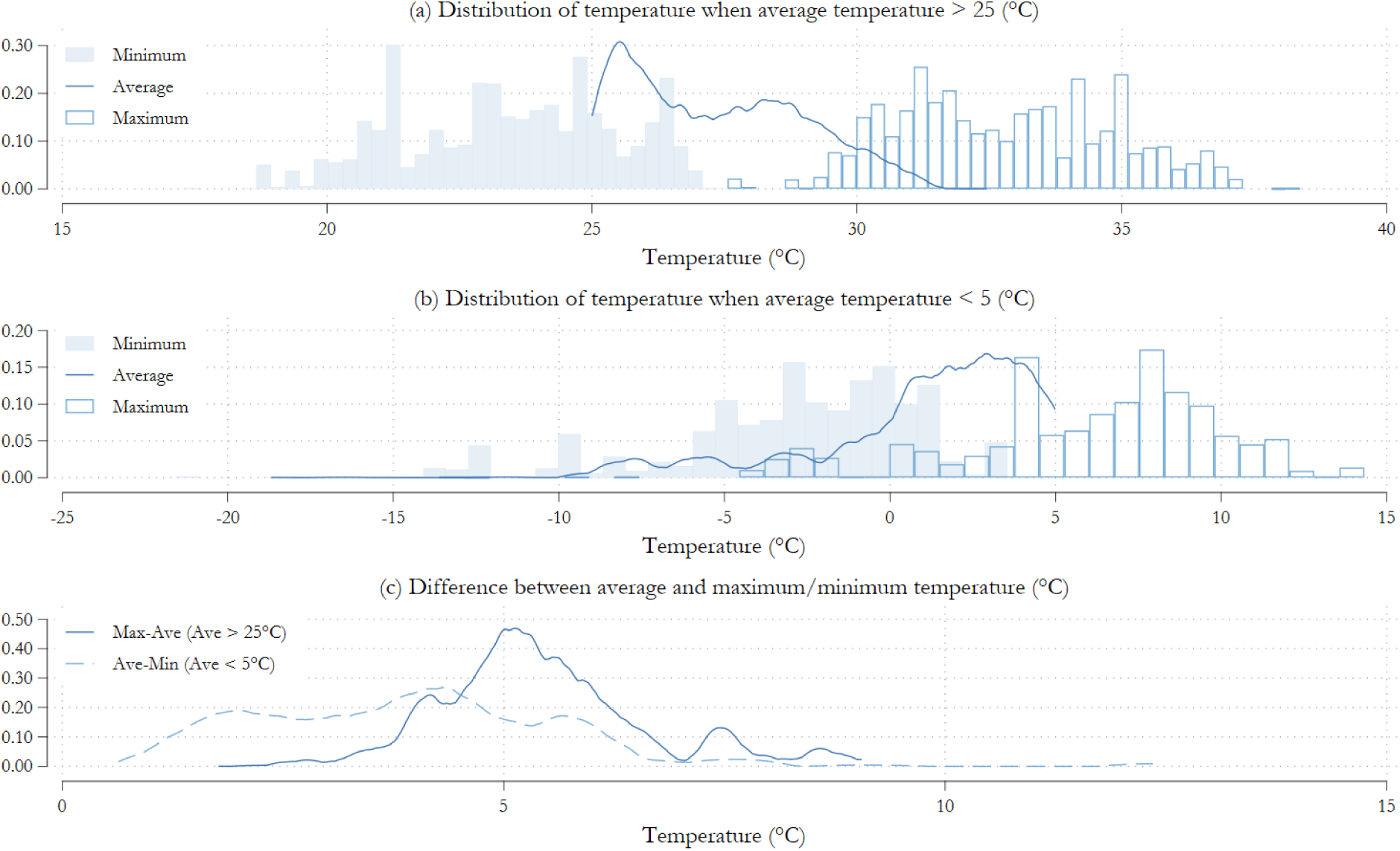
Summary of temperature variables in the research period. *Notes:* These figures provide an additional summary of temperature variables for the research period. Figure (a) shows the distribution of temperature when the average temperature is > 25 (°C); Figure (b) shows the distribution of temperature when the average temperature is < 5 (°C); Figure (c) shows the difference between the average and maximum/minimum temperature (°C).

**Figure A5.**
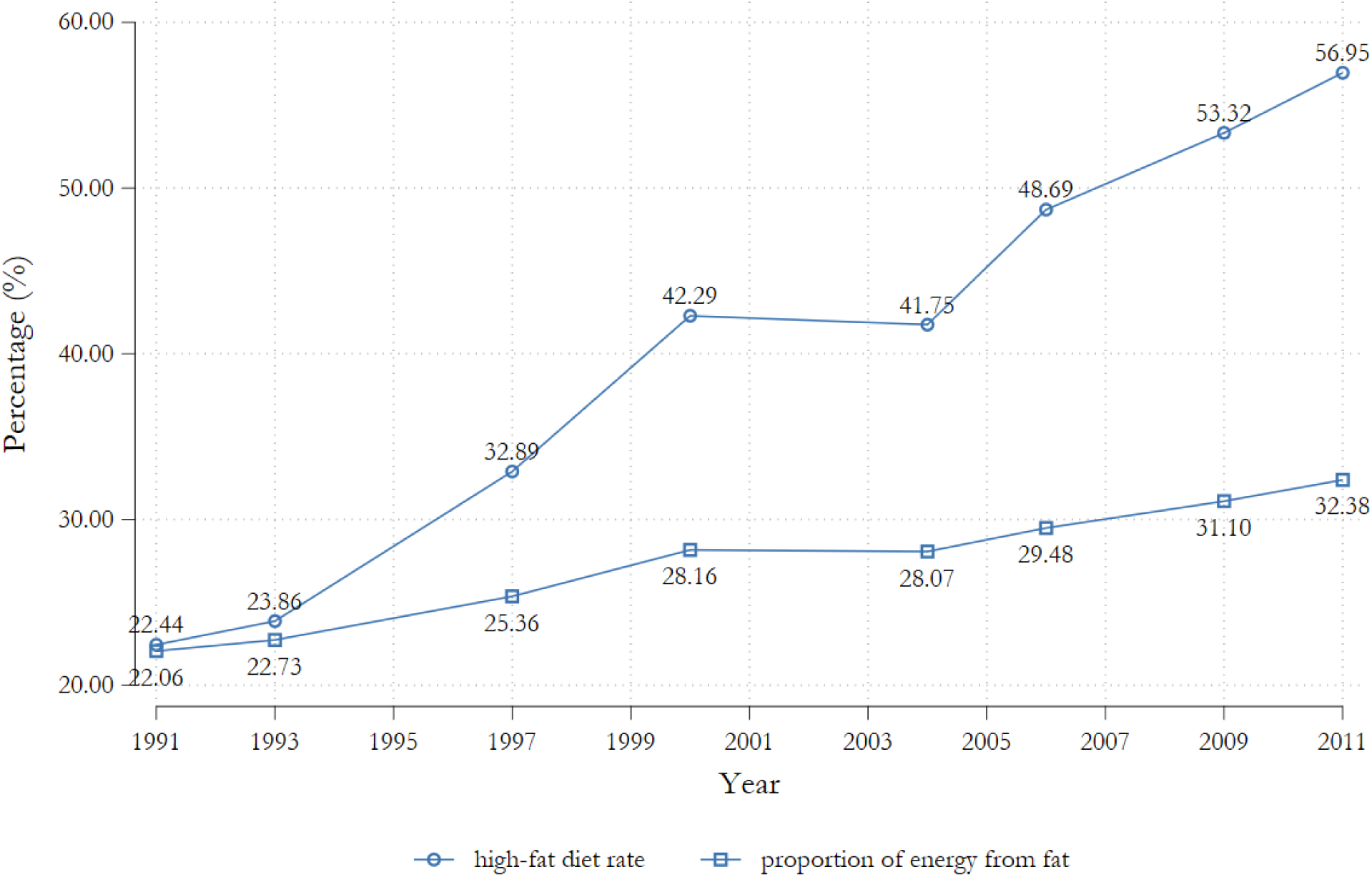
**Trends of energy intake from fat and high-fat diet rate (%)***Notes:* This figure shows the trends of energy intake from fat and high-fat diet rate (%) in the CHNS survey sample from 1991 to 2011.

**Figure A6.**
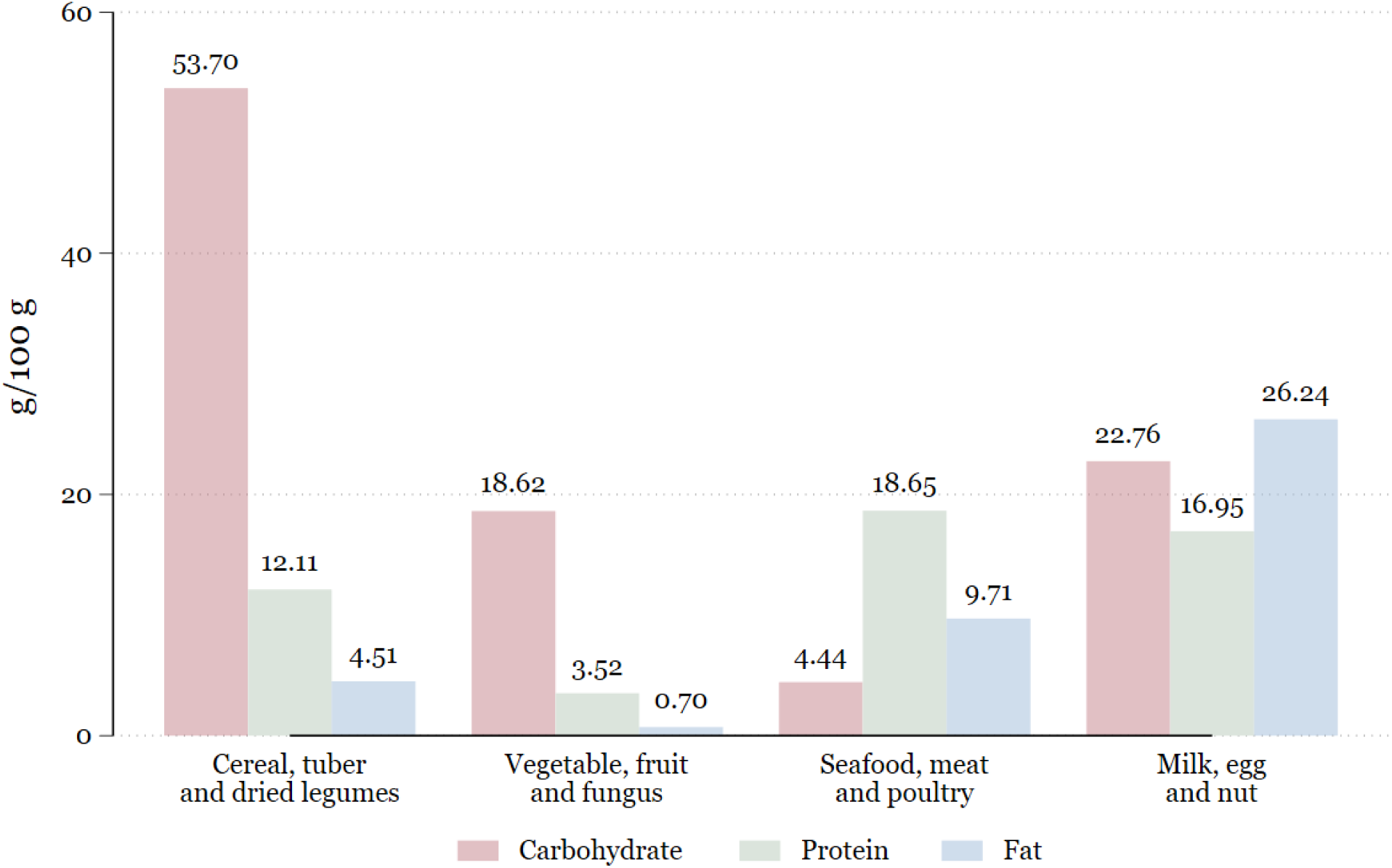
Contribution of the four major food groups to macronutrients. *Notes:* This figure shows the nutrient density of the four major food groups. The calculations rely on nutritional composition data provided by China’s Nutrition of Main Foods (China CDC, 2018). Each bar represents the average gram of carbohydrate, protein, or fat intake derived from 100g of food in the corresponding group.

**Figure A7.**
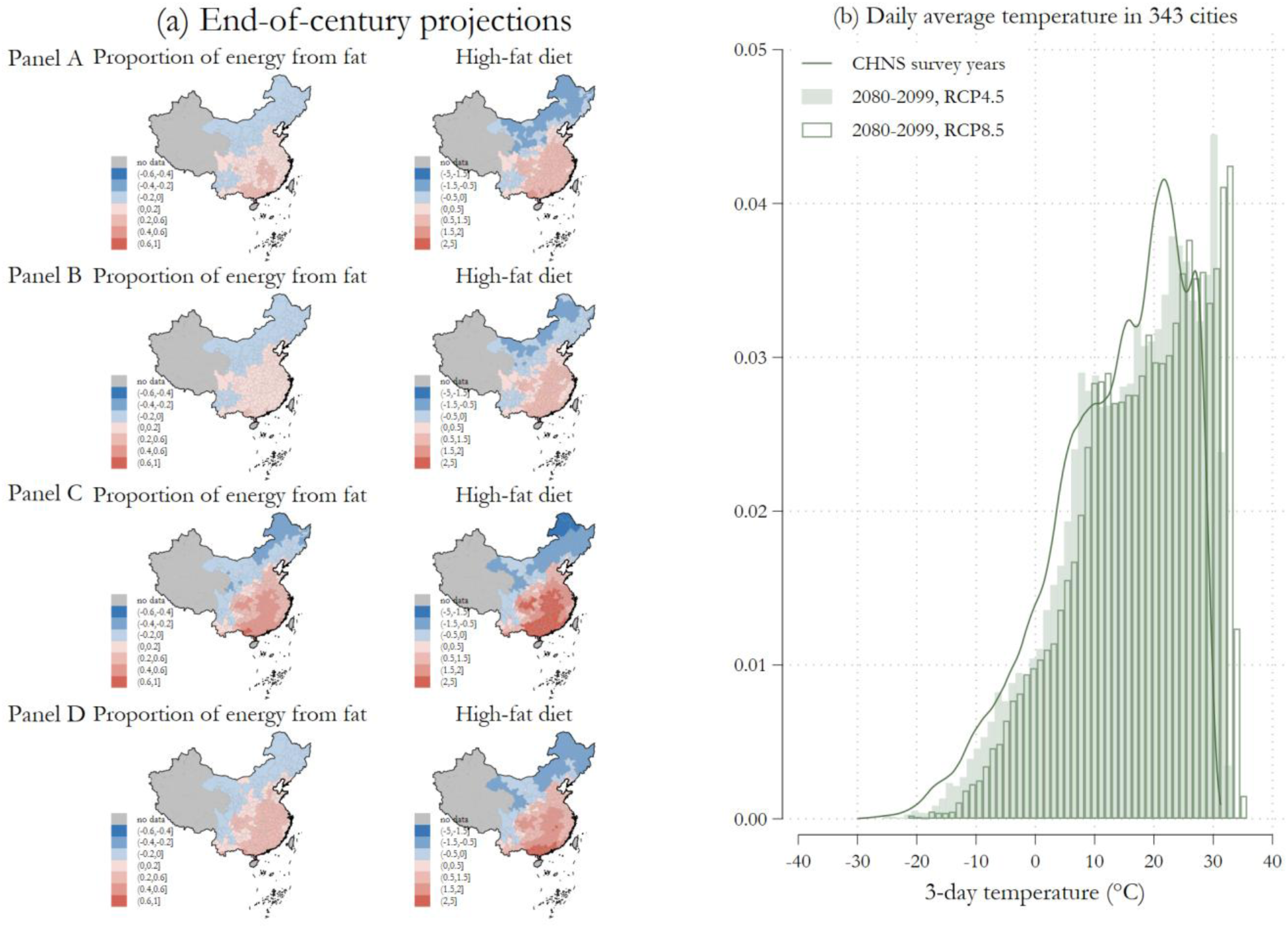
End-of-century projections and distribution of daily average temperature in 343 cities. *Notes:* Figure (a) shows the predicted impact of climate change on the proportion of energy from fat and the probability of a high-fat diet for each city in 2080-2099 relative to CHNS survey waves. Due to the limited coverage of provinces of CHNS (see Figure A1), results based on this database may not be representative of the national average, especially in the Northwest region and the Tibetan Plateau. Therefore, we remove Qinghai, Tibet, and Xinjiang provinces from the forecast. Panel A and Panel C show the projection results based on equation (4) under the climate scenarios RCP4.5 and RCP8.5, respectively. Panel B and Panel D show the projection results based on equation (5), which considers the adaptation effects of the fans, heating systems, and refrigerators under the climate scenarios RCP4.5 and RCP8.5. All the scales in subgraphs are uniform. Red indicates an increase in the proportion or probability, while blue shows the opposite effect. Figure (b) shows the distribution of daily temperature in three scenarios. The first scenario is the average daily temperature during the CHNS survey years, which are 1991, 1993, 1997, 2000, 2004, 2006, 2009, and 2011. The second scenario is the projected average daily temperature in RCP4.5, and the third is RCP8.5. We average the data to the city-day level and plot the density picture.

## Appendix Tables

**Table A1.** Examples of the effect of temperature on agriculture in the literature

| Article | Area | Time period | Results |
| --- | --- | --- | --- |
| Burke and Emerick (2016) | U.S. | 1980-2000 | (1) Each extra degree-day of heat above 29°C leads to a 0.44% reduction in total corn yield. |
| Chen et al. (2016) | China | 2000-2009 | (1) Crop yields have nonlinear and inverted U-shaped relationships with weather variables.<br>(2) Over the research period, global warming has led to an economic loss of approximately \$820 million for China's corn and soybean sectors. |
| Aragón et al. (2021) | Peru | 2007-2015 | (1) Extreme temperatures are associated with reductions in agricultural productivity, but an increase in area planted, which means farmers partially offset the decline in output by increasing the area planted.<br>(2) Each additional average cumulative exposure to temperatures above 33°C during the growing season results in a 7% decrease in agricultural productivity and a 6% increase in planting area. |
| Chen and Gong (2021) | China | 1981-2015 | (1) In the short run, an additional one-day cumulative exposure to temperatures above 33°C during the whole year leads to a 2.6% decrease in agricultural TFP and a 4.4% decrease in yield.<br>(2) The long-run impact of extreme heating on TFP and agricultural yield is about 1.6% and 2.3%, respectively. |
| Miller et al. (2021) | Global | 1979-2016 | (1) An abnormally hot day (defined as a cell-level median (mean) of 89 (86) °F) not far from being preceded by at least eight others reduces per-capita agricultural value added by approximately 1.7%. |
| Chen et al. (2023) | China | 1995-2015 | (1) An additional one-day exposure to temperatures above 35°C would lower the aggregate output of northern agriculture by about 4.95%, relative to the reference bin (10-15°C).<br>(2) The reduction in aggregate output is mostly driven by the cropping, livestock, and fishery sectors, with output value losses of 2.58%, 2.61%, and 2.11%, respectively.<br>(3) There's no significant impact of temperature on agricultural output in southern China. |
| Wang et al. (2024) | China | 1981-2010 | (1) During 1981-1995, an additional 100-degree-day exposure to temperatures above 28°C led to a 23% loss in corn yield, while the corresponding estimate in 1996-2010 is -11.5%.<br>(2) During 1981-1995, an additional 100-degree-day exposure to temperatures above 26°C led to a 15.7% loss in soybean yield, while the corresponding estimate in 1996-2010 is -8.3%.<br>(3) The overall impacts of 100-day exposure to extreme high temperatures on corn and soybean yields from 1996 to 2010 are |
|  |  |  | 40% to 50% less than those from 1981 to 1995. This results in a loss reduction by about 14.5% of the national corn production and 7% of the national soybean production. |
| Cui and Zhong (2024) | China | 1981-2015 | (1) In cold regions where the average temperature in 1980 was below 10°C, a 0.1°C increase in the 30-year average temperature leads to a 2.4% expansion in cropland.<br>(2) In mild regions, where the initial temperature ranges between 10-20°C, a 0.1°C long-term warming decreases total cropland areas by 1.2%.<br>(3) This negative impact in hot regions is about 1.9% per 0.1°C. |
| Cui and Tang (2024) | Northeast<br>China | 2004-2014 | (1) An additional degree-day accumulated above 32°C in the growing season (i.e., April to October) reduces corn yields by approximately 5.4% and household total corn outputs by about 5.0%.<br>(2) This also leads to a reduction in aggregate farm revenue and net household income for corn households by 2.9% and 2.1%, respectively. |

*Notes:* This table shows some of the results about the effects of temperatures on agriculture in the literature.

**Table A2.** Descriptive statistics

| Variables | Unit | Number | Mean | SD | Min | Max |
| --- | --- | --- | --- | --- | --- | --- |
| Total consumption | 100g/3-day | 91,570 | 30.58 | 10.67 | 6.750 | 72.73 |
| Energy | 100kcal/3-day | 92,500 | 63.98 | 20.20 | 12.34 | 155.2 |
| Carbohydrate | 100g/3-day | 93,899 | 9.705 | 3.764 | 2.151 | 23.78 |
| Protein | 100g/3-day | 93,900 | 1.953 | 0.697 | 0.451 | 4.581 |
| Fat | 100g/3-day | 93,899 | 1.929 | 1.056 | 0.156 | 6.365 |
| Proportion of energy from fat | - | 92,500 | 0.272 | 0.115 | 0.0181 | 0.780 |
| High-fat diet | - | 92,500 | 0.395 | 0.489 | 0 | 1 |
| Cereal, tuber, and dried legumes | 100g/3-day | 91,570 | 15.54 | 6.580 | 0 | 60.50 |
| Vegetable, fruit, and fungus | 100g/3-day | 91,570 | 10.85 | 6.076 | 0 | 55.70 |
| Seafood, meat, and poultry | 100g/3-day | 91,570 | 3.110 | 3.146 | 0 | 34.03 |
| Milk, egg, and nut | 100g/3-day | 91,570 | 1.072 | 1.810 | 0 | 21.90 |
| Sugar and salt | 100g/3-day | 64,627 | 0.328 | 0.255 | 0 | 2.946 |
| Oil | 100g/3-day | 64,924 | 1.137 | 0.774 | 0 | 6 |
| BMI | - | 68,933 | 22.75 | 3.287 | 14.79 | 39.37 |
| Urban | - | 102,089 | 0.292 | 0.455 | 0 | 1 |
| Age | Year | 102,931 | 36.85 | 20.91 | 0 | 101 |
| Male | - | 102,942 | 0.497 | 0.500 | 0 | 1 |
| Air conditioner | - | 102,942 | 0.137 | 0.344 | 0 | 1 |
| Fan | - | 102,942 | 0.749 | 0.434 | 0 | 1 |
| Refrigerator | - | 102,942 | 0.403 | 0.490 | 0 | 1 |
| Heating system | - | 102,942 | 0.0799 | 0.271 | 0 | 1 |
| 3-day CDD | degree-days | 102,942 | 0.668 | 2.433 | 0 | 20.42 |
| 3-day HDD | degree-days | 102,942 | 0.504 | 2.913 | 0 | 45.79 |
| 3-day average humidity | % | 102,942 | 73.39 | 11.09 | 31.76 | 98.12 |
| 3-day average precipitation | mm | 102,942 | 1.948 | 3.874 | 0 | 33.16 |
| 3-day average wind speed | m/s | 102,942 | 1.838 | 0.843 | 0.140 | 6.630 |
| 3-day average sunshine duration | h | 102,942 | 5.356 | 3.104 | 0 | 12.09 |

*Notes:* This table shows the statistics of key variables in the empirical analysis. The variables related to food and macronutrients calculate the total consumption in the past 3 days, and the units are in 100g (or 100 kcal).

**Table A3.** Baseline results

| Variables | (1)<br>Proportion<br>of energy<br>from fat | (2)<br>High-fat diet | (3)<br>Total<br>consumption | (4)<br>Energy | (5)<br>Carbohydrate | (6)<br>Protein | (7)<br>Fat |
| --- | --- | --- | --- | --- | --- | --- | --- |
| 3-day CDD | 0.0807***<br>(0.0281) | 0.4039***<br>(0.1356) | -0.0284<br>(0.0959) | -0.1640**<br>(0.0820) | -0.2387***<br>(0.0913) | -0.3484***<br>(0.0932) | 0.1514<br>(0.1526) |
| 3-day HDD | 0.0382*<br>(0.0207) | 0.1951*<br>(0.1017) | 0.1588**<br>(0.0748) | 0.1264*<br>(0.0646) | 0.0566<br>(0.0719) | 0.1577**<br>(0.0710) | 0.3648***<br>(0.1058) |
| Observations | 86,147 | 86,147 | 85,087 | 86,147 | 87,612 | 87,593 | 87,584 |
| R-squared | 0.5429 | 0.4409 | 0.4789 | 0.5197 | 0.5845 | 0.4929 | 0.4855 |
| Dep. Var. Mean | 0.272 | 0.395 | 30.58 | 63.98 | 9.705 | 1.953 | 1.929 |
| Individual FE | YES | YES | YES | YES | YES | YES | YES |
| Year-month FE | YES | YES | YES | YES | YES | YES | YES |
| Province-year FE | YES | YES | YES | YES | YES | YES | YES |
| Province-month FE | YES | YES | YES | YES | YES | YES | YES |

**Table A4.** Wald tests for the equality of the fat coefficient across macronutrients

|  | Fat with Carbohydrate | Fat with Protein |
| --- | --- | --- |
| Wald test, $\beta_{\text{Fat}} = \beta_j$ under heat (p) | 0.008 | 0.000 |
| Wald test, $\beta_{\text{Fat}} = \beta_j$ under cold (p) | 0.010 | 0.046 |
| Observations | 86,147 | 86,147 |

*Notes:* Each entry is the p-value from a Wald test of the null hypothesis that the temperature coefficient on fat equals that on the indicated macronutrient (carbohydrate or protein) within the same temperature regime; the two rows correspond to the 3-day CDD (heat) and 3-day HDD (cold) responses. The tests come from a single stacked regression of the three log macronutrient outcomes (log carbohydrate, log protein, and log fat intake per 100 g over 3 days), in which the two temperature variables, the weather controls, and all fixed effects are fully interacted with a macronutrient indicator. This specification numerically reproduces the coefficients of the three separate baseline regressions in Table A3 while delivering a joint covariance matrix across the three equations, and it is equivalent to seemingly unrelated regressions that accommodate high-dimensional fixed effects. Fixed effects are individual, year-by-month, province-by-year, and province-by-month, each interacted with the macronutrient indicator, and standard errors are clustered at the individual level. The joint null that the fat coefficient equals both the carbohydrate and protein coefficients is also rejected, under heat ( $F(2, 21177) = 8.43$ , $p = 0.000$ ) and under cold ( $F(2, 21177) = 3.60$ , $p = 0.027$ ). The estimation sample comprises observations with all three macronutrients non-missing (86,147 individual-wave observations per equation).

**Table A5.** Suggestive evidence on the correlation between BMI and fat intake

|  | (1) | (2) | (3) | (4) | (5) | (6) |
| --- | --- | --- | --- | --- | --- | --- |
| Variables | BMI | BMI | BMI | Overweight | Overweight | Overweight |
| Fat | 0.3662***<br>(0.0431) |  |  | 0.0508***<br>(0.0064) |  |  |
| Proportion of energy from fat |  | 2.6835***<br>(0.3329) |  |  | 0.3672***<br>(0.0488) |  |
| High-fat diet |  |  | 0.2886***<br>(0.0563) |  |  | 0.0414***<br>(0.0083) |
| Energy | 0.0076***<br>(0.0023) | 0.0189***<br>(0.0018) | 0.0192***<br>(0.0018) | 0.0002<br>(0.0003) | 0.0018***<br>(0.0003) | 0.0018***<br>(0.0003) |
| Observations | 18,662 | 18,662 | 18,662 | 18,662 | 18,662 | 18,662 |
| R-squared | 0.0979 | 0.0976 | 0.0958 | 0.0684 | 0.0682 | 0.0666 |
| Control | NO | NO | NO | NO | NO | NO |
| Community FE | YES | YES | YES | YES | YES | YES |
|  | (7) | (8) | (9) | (10) | (11) | (12) |
| Variables | BMI | BMI | BMI | Overweight | Overweight | Overweight |
| Fat | 0.2228***<br>(0.0587) |  |  | 0.0327***<br>(0.0089) |  |  |
| Proportion of energy from fat |  | 1.6244***<br>(0.4550) |  |  | 0.2396***<br>(0.0683) |  |
| High-fat diet |  |  | 0.1493**<br>(0.0701) |  |  | 0.0262**<br>(0.0106) |
| Energy | 0.0170***<br>(0.0034) | 0.0241***<br>(0.0027) | 0.0248***<br>(0.0026) | 0.0014***<br>(0.0005) | 0.0024***<br>(0.0004) | 0.0025***<br>(0.0004) |
| Observations | 12,120 | 12,120 | 12,120 | 12,120 | 12,120 | 12,120 |
| R-squared | 0.1029 | 0.1028 | 0.1022 | 0.0740 | 0.0739 | 0.0734 |
| Control | YES | YES | YES | YES | YES | YES |
| Community FE | YES | YES | YES | YES | YES | YES |

Notes: (1) We use the BMI of adults (age $\geq 18$ ) as the outcome variable. (2) Columns (1)-(6) do not include any control variables. Columns (7)-(12) control for the averages of working hours and physical activity variables across all survey waves and gender. (3) Community fixed effects are involved in all regressions. (4) Standard errors shown in parentheses are clustered at the individual level. (5) \* $p < 0.1$ , \*\* $p < 0.05$ , \*\*\* $p < 0.01$ .

**Table A6.**
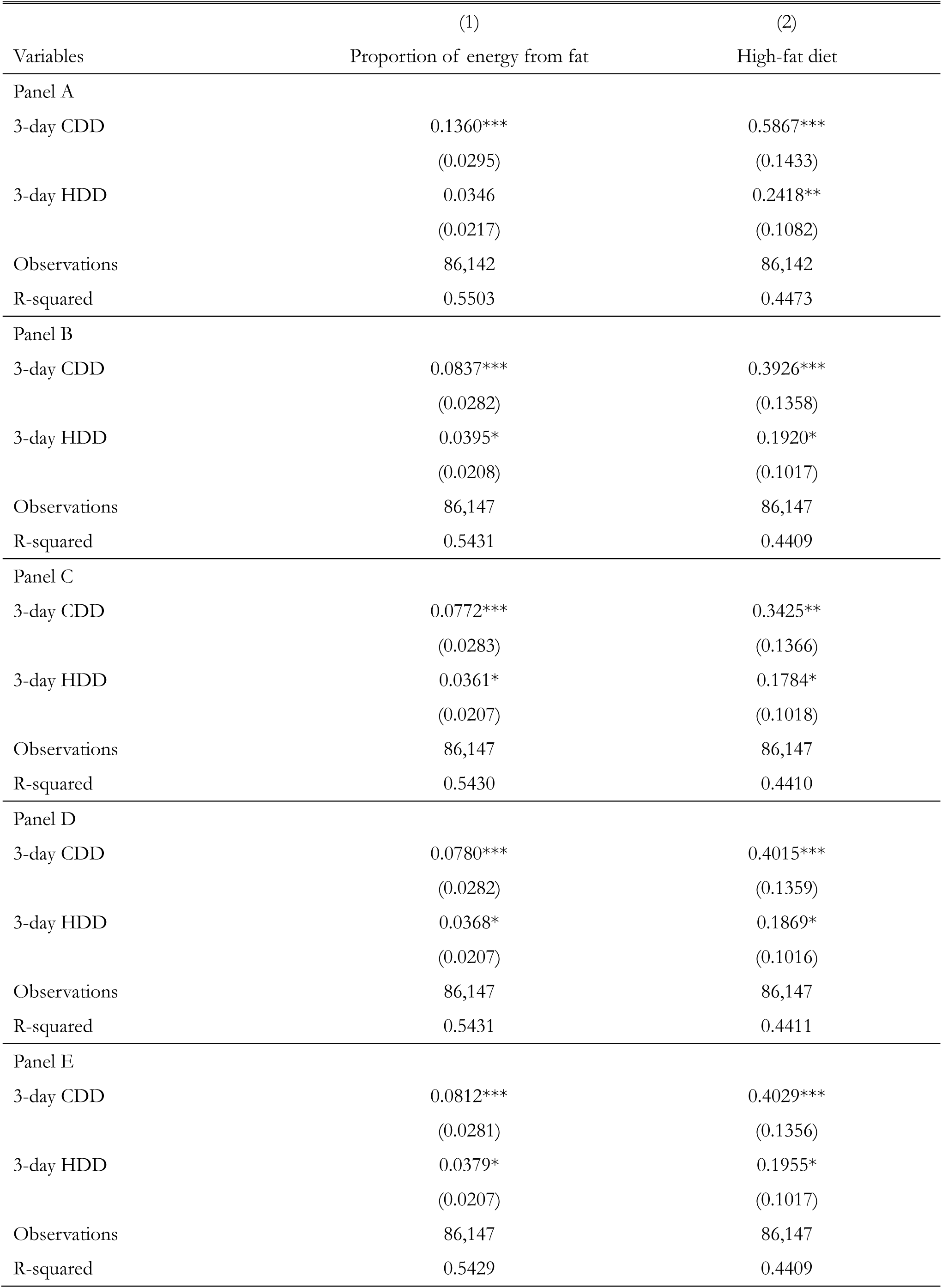
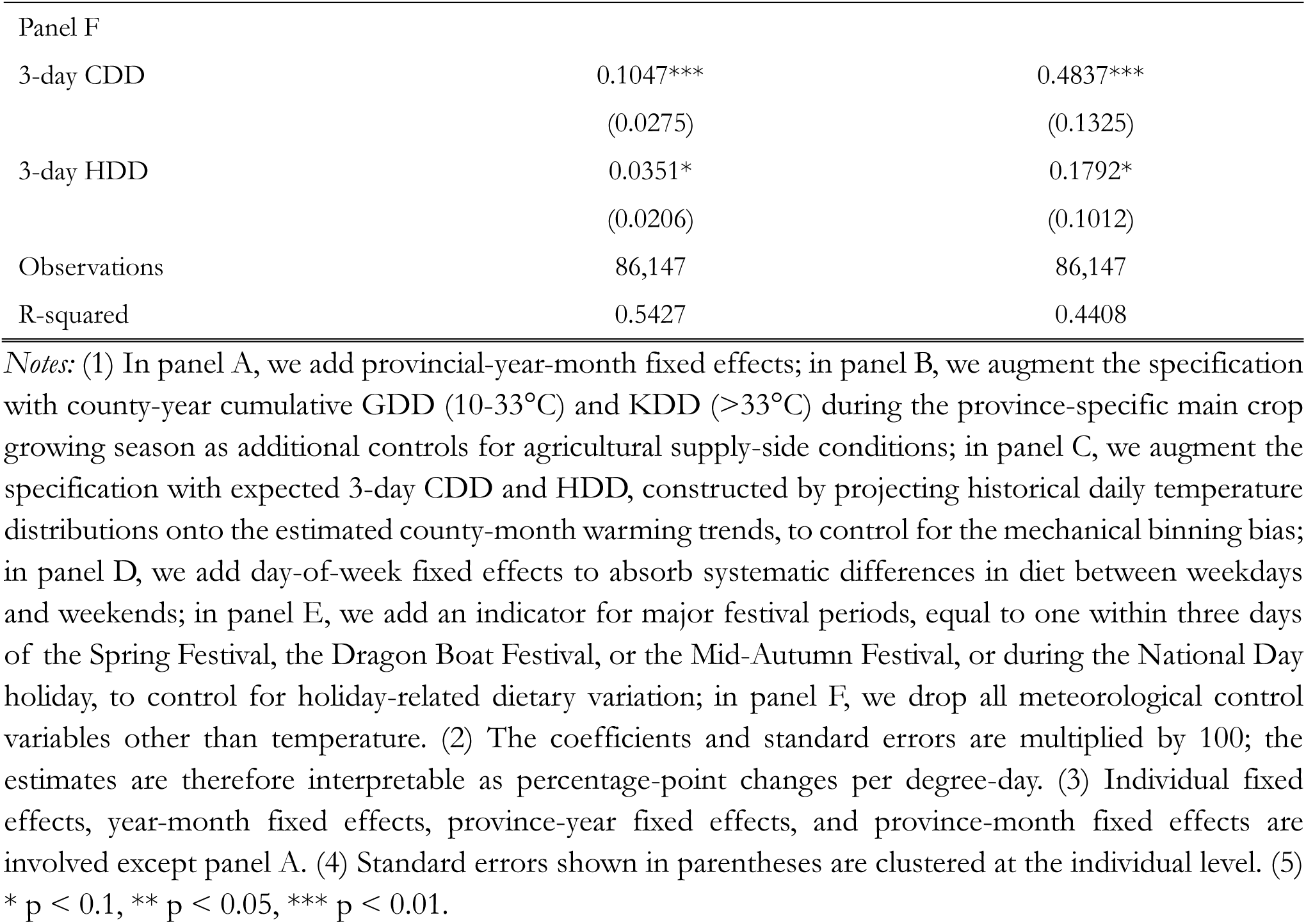
Robustness tests

| Variables | (1)<br>Proportion of energy from fat | (2)<br>High-fat diet |
| --- | --- | --- |
| Panel A |  |  |
| 3-day CDD | 0.1360***<br>(0.0295) | 0.5867***<br>(0.1433) |
| 3-day HDD | 0.0346<br>(0.0217) | 0.2418**<br>(0.1082) |
| Observations | 86,142 | 86,142 |
| R-squared | 0.5503 | 0.4473 |
| Panel B |  |  |
| 3-day CDD | 0.0837***<br>(0.0282) | 0.3926***<br>(0.1358) |
| 3-day HDD | 0.0395*<br>(0.0208) | 0.1920*<br>(0.1017) |
| Observations | 86,147 | 86,147 |
| R-squared | 0.5431 | 0.4409 |
| Panel C |  |  |
| 3-day CDD | 0.0772***<br>(0.0283) | 0.3425**<br>(0.1366) |
| 3-day HDD | 0.0361*<br>(0.0207) | 0.1784*<br>(0.1018) |
| Observations | 86,147 | 86,147 |
| R-squared | 0.5430 | 0.4410 |
| Panel D |  |  |
| 3-day CDD | 0.0780***<br>(0.0282) | 0.4015***<br>(0.1359) |
| 3-day HDD | 0.0368*<br>(0.0207) | 0.1869*<br>(0.1016) |
| Observations | 86,147 | 86,147 |
| R-squared | 0.5431 | 0.4411 |
| Panel E |  |  |
| 3-day CDD | 0.0812***<br>(0.0281) | 0.4029***<br>(0.1356) |
| 3-day HDD | 0.0379*<br>(0.0207) | 0.1955*<br>(0.1017) |
| Observations | 86,147 | 86,147 |
| R-squared | 0.5429 | 0.4409 |
| Panel F |  |  |
| 3-day CDD | 0.1047*** | 0.4837*** |
|  | (0.0275) | (0.1325) |
| 3-day HDD | 0.0351* | 0.1792* |
|  | (0.0206) | (0.1012) |
| Observations | 86,147 | 86,147 |
| R-squared | 0.5427 | 0.4408 |

*Notes:* (1) In panel A, we add provincial-year-month fixed effects; in panel B, we augment the specification with county-year cumulative GDD (10-33°C) and KDD (>33°C) during the province-specific main crop growing season as additional controls for agricultural supply-side conditions; in panel C, we augment the specification with expected 3-day CDD and HDD, constructed by projecting historical daily temperature distributions onto the estimated county-month warming trends, to control for the mechanical binning bias; in panel D, we add day-of-week fixed effects to absorb systematic differences in diet between weekdays and weekends; in panel E, we add an indicator for major festival periods, equal to one within three days of the Spring Festival, the Dragon Boat Festival, or the Mid-Autumn Festival, or during the National Day holiday, to control for holiday-related dietary variation; in panel F, we drop all meteorological control variables other than temperature. (2) The coefficients and standard errors are multiplied by 100; the estimates are therefore interpretable as percentage-point changes per degree-day. (3) Individual fixed effects, year-month fixed effects, province-year fixed effects, and province-month fixed effects are involved except panel A. (4) Standard errors shown in parentheses are clustered at the individual level. (5) \* $p < 0.1$ , \*\* $p < 0.05$ , \*\*\* $p < 0.01$ .

**Table A7.** Robustness tests – different temperature definition

| Variables | (1)<br>Proportion of energy from fat | (2)<br>High-fat diet |
| --- | --- | --- |
| Cutoff: Ave-temp 4-26°C |  |  |
| 3-day CDD | 0.1524***<br>(0.0355) | 0.6798***<br>(0.1709) |
| 3-day HDD | 0.0467*<br>(0.0238) | 0.2383**<br>(0.1165) |
| Observations | 86,147 | 86,147 |
| R-squared | 0.5429 | 0.4409 |
| Cutoff: Ave-temp 3-27°C |  |  |
| 3-day CDD | 0.2354***<br>(0.0461) | 1.0426***<br>(0.2229) |
| 3-day HDD | 0.0574**<br>(0.0274) | 0.2969**<br>(0.1342) |
| Observations | 86,147 | 86,147 |
| R-squared | 0.5430 | 0.4410 |
| Cutoff: Ave-temp 2-28°C |  |  |
| 3-day CDD | 0.2761***<br>(0.0621) | 1.4002***<br>(0.3021) |
| 3-day HDD | 0.0667**<br>(0.0320) | 0.3730**<br>(0.1568) |
| Observations | 86,147 | 86,147 |
| R-squared | 0.5430 | 0.4410 |
| Cutoff: Max-temp 30°C, min-temp 0°C |  |  |
| 3-day CDD | 0.0822***<br>(0.0211) | 0.3343***<br>(0.0997) |
| 3-day HDD | 0.0330<br>(0.0225) | 0.1890*<br>(0.1121) |
| Observations | 86,147 | 86,147 |
| R-squared | 0.5429 | 0.4409 |

**Table A8.** Different foods

| Variables | (1) | (2) | (3) |
| --- | --- | --- | --- |
|  | Cereal, tuber, and dried legumes | Vegetable, fruit, and fungus | Seafood, meat, and poultry |
| 3-day CDD | -0.0007<br>(0.0140) | 0.0365**<br>(0.0183) | -0.0152**<br>(0.0076) |
| 3-day HDD | 0.0063<br>(0.0133) | 0.0272**<br>(0.0127) | 0.0037<br>(0.0048) |
| Observations | 83,085 | 83,085 | 83,085 |
| R-squared | 0.5367 | 0.4031 | 0.5835 |
| Dep. Var. Mean | 15.54 | 10.85 | 3.110 |
|  | (4) | (5) | (6) |
|  | Milk, egg, and nut | Sugar and salt | Oil |
| 3-day CDD | 0.0096**<br>(0.0041) | 0.0010<br>(0.0007) | -0.0042*<br>(0.0023) |
| 3-day HDD | 0.0154***<br>(0.0052) | -0.0011*<br>(0.0006) | 0.0000<br>(0.0021) |
| Observations | 83,085 | 56,620 | 56,963 |
| R-squared | 0.5336 | 0.4512 | 0.4095 |
| Dep. Var. Mean | 1.072 | 0.328 | 1.137 |
| Individual FE | YES | YES | YES |
| Year-month FE | YES | YES | YES |
| Province-year FE | YES | YES | YES |
| Province-month FE | YES | YES | YES |

*Notes:* (1) We use the original levels as dependent variables in the regression because these dependent variables contain zero observations. (2) Individual fixed effects, year-month fixed effects, province-year fixed effects, and province-month fixed effects are involved in all regressions. (3) Standard errors shown in parentheses are clustered at the individual level. (4) \* $p < 0.1$ , \*\* $p < 0.05$ , \*\*\* $p < 0.01$ . (5) Dep. Var. Mean shows the mean of Y, which is the original levels of each type of food in total food consumption, measured in 100g/3-day.

**Table A9.**
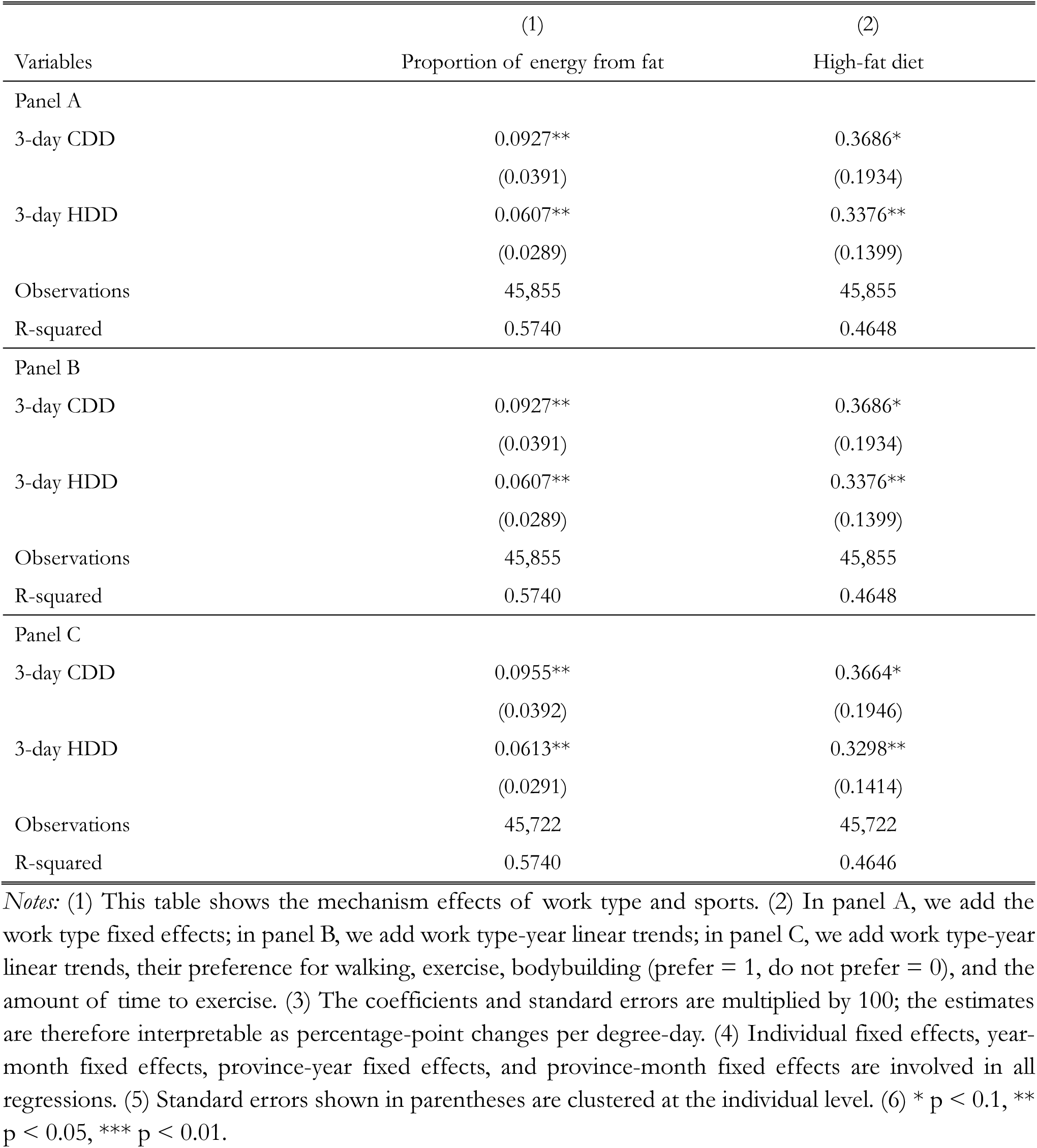
Mechanism effects – work type and sports

| Variables | (1)<br>Proportion of energy from fat | (2)<br>High-fat diet |
| --- | --- | --- |
| Panel A |  |  |
| 3-day CDD | 0.0927**<br>(0.0391) | 0.3686*<br>(0.1934) |
| 3-day HDD | 0.0607**<br>(0.0289) | 0.3376**<br>(0.1399) |
| Observations | 45,855 | 45,855 |
| R-squared | 0.5740 | 0.4648 |
| Panel B |  |  |
| 3-day CDD | 0.0927**<br>(0.0391) | 0.3686*<br>(0.1934) |
| 3-day HDD | 0.0607**<br>(0.0289) | 0.3376**<br>(0.1399) |
| Observations | 45,855 | 45,855 |
| R-squared | 0.5740 | 0.4648 |
| Panel C |  |  |
| 3-day CDD | 0.0955**<br>(0.0392) | 0.3664*<br>(0.1946) |
| 3-day HDD | 0.0613**<br>(0.0291) | 0.3298**<br>(0.1414) |
| Observations | 45,722 | 45,722 |
| R-squared | 0.5740 | 0.4646 |

*Notes:* (1) This table shows the mechanism effects of work type and sports. (2) In panel A, we add the work type fixed effects; in panel B, we add work type-year linear trends; in panel C, we add work type-year linear trends, their preference for walking, exercise, bodybuilding (prefer = 1, do not prefer = 0), and the amount of time to exercise. (3) The coefficients and standard errors are multiplied by 100; the estimates are therefore interpretable as percentage-point changes per degree-day. (4) Individual fixed effects, year-month fixed effects, province-year fixed effects, and province-month fixed effects are involved in all regressions. (5) Standard errors shown in parentheses are clustered at the individual level. (6) \* $p < 0.1$ , \*\* $p < 0.05$ , \*\*\* $p < 0.01$ .

**Table A10.** Mechanism effects – work hours

| Variables | (1)<br>Last week's work hours | (2)<br>Last week's work hours |
| --- | --- | --- |
| Last week's average temperature | 0.0016<br>(0.0086) |  |
| Last week CDD |  | -0.0661*<br>(0.0402) |
| Last week HDD |  | -0.0764**<br>(0.0336) |
| Observations | 27,295 | 27,295 |
| R-squared | 0.5250 | 0.5252 |
| Dep. Var. Mean | 42.06 | 42.06 |
| Individual FE | YES | YES |
| Year-month FE | YES | YES |
| Province-year FE | YES | YES |
| Province-month FE | YES | YES |

*Notes:* (1) This table shows the mechanism effects of work time. (2) Since the outcome variable considers the work hours in the last week, we calculate the last week's average temperature, CDD, and HDD for each individual. In column (1), the key independent variable is the last week's average temperature; in column (2), the key independent variables are last week's CDD and HDD. (3) Individual fixed effects, year-month fixed effects, province-year fixed effects, and province-month fixed effects are involved in all regressions. (4) Standard errors shown in parentheses are clustered at the individual level. (5) \* $p < 0.1$ , \*\* $p < 0.05$ , \*\*\* $p < 0.01$ .

**Table A11.** Heterogeneous effects

| Variables | (1)<br>Proportion of energy from fat | (2)<br>High-fat diet |
| --- | --- | --- |
| Panel A – Age |  |  |
| 3-day CDD * children | 0.0597<br>(0.0488) | 0.2293<br>(0.2244) |
| 3-day CDD * adult | 0.1129***<br>(0.0309) | 0.5205***<br>(0.1501) |
| 3-day CDD * elder | 0.0074<br>(0.0441) | 0.2158<br>(0.2117) |
| 3-day HDD * children | 0.1089***<br>(0.0402) | 0.5999***<br>(0.2013) |
| 3-day HDD * adult | 0.0354<br>(0.0231) | 0.1990*<br>(0.1123) |
| 3-day HDD * elder | -0.0049<br>(0.0360) | -0.1194<br>(0.1849) |
| Observations | 86,145 | 86,145 |
| R-squared | 0.5430 | 0.4410 |
| Panel B – Gender |  |  |
| 3-day CDD * female | 0.0833**<br>(0.0333) | 0.4601***<br>(0.1593) |
| 3-day CDD * male | 0.0781**<br>(0.0325) | 0.3485**<br>(0.1571) |
| 3-day HDD * female | 0.0517**<br>(0.0254) | 0.1747<br>(0.1210) |
| 3-day HDD * male | 0.0235<br>(0.0250) | 0.2172*<br>(0.1232) |
| Observations | 86,147 | 86,147 |
| R-squared | 0.5429 | 0.4409 |
| Panel C – Rural and urban |  |  |
| 3-day CDD * rural | 0.0947***<br>(0.0294) | 0.4339***<br>(0.1418) |
| 3-day CDD * urban | -0.0282<br>(0.0445) | 0.1434<br>(0.2195) |
| 3-day HDD * rural | 0.0931***<br>(0.0260) | 0.4475***<br>(0.1238) |
| 3-day HDD * urban | -0.0280<br>(0.0260) | -0.1228<br>(0.1297) |
| Observations | 85,345 | 85,345 |
| R-squared | 0.5441 | 0.4422 |
| Panel D – Wealth |  |  |
| 3-day CDD * poor | 0.1095***<br>(0.0317) | 0.5135***<br>(0.1524) |
| 3-day CDD * rich | 0.0329 | 0.2194 |
|  | (0.0344) | (0.1639) |
| 3-day HDD * poor | 0.0652** | 0.2532* |
|  | (0.0266) | (0.1301) |
| 3-day HDD * rich | 0.0233 | 0.1758 |
|  | (0.0248) | (0.1222) |
| Observations | 86,147 | 86,147 |
| R-squared | 0.5434 | 0.4414 |

*Notes:* (1) Each panel shows the regression results of different heterogeneity analyses. (2) We use the original levels of the proportion of energy from fat and the high-fat diet index, because they already represent the meaning of proportion. (3) The coefficients and standard errors are multiplied by 100; the estimates are therefore interpretable as percentage-point changes per degree-day. (4) Individual fixed effects, year-month fixed effects, province-year fixed effects, and province-month fixed effects are involved in all regressions. (5) Standard errors shown in parentheses are clustered at the individual level. (6) \* $p < 0.1$ , \*\* $p < 0.05$ , \*\*\* $p < 0.01$ .

**Table A12.** Descriptive statistics in rural, urban, and different wealth groups

| Variables | Unit | Rural | Urban | diff | t-value |
| --- | --- | --- | --- | --- | --- |
| Total consumption | 100g/3-day | 30.82 | 30.09 | 0.729 | 9.425*** |
| Energy | 100kcal/3-day | 64.80 | 62.16 | 2.643 | 18.179*** |
| Carbohydrate | 100g/3-day | 10.16 | 8.681 | 1.475 | 55.594*** |
| Protein | 100g/3-day | 1.937 | 1.994 | -0.0580 | -11.574*** |
| Fat | 100g/3-day | 1.828 | 2.163 | -0.336 | -44.825*** |
| Proportion of energy from fat | - | 0.255 | 0.311 | -0.0560 | -68.852*** |
| High-fat diet | - | 0.334 | 0.532 | -0.198 | -57.249*** |
| Cereal, tuber and dried legumes | 100g/3-day | 16.18 | 14.09 | 2.095 | 44.327*** |
| Vegetable, fruit and fungus | 100g/3-day | 11.09 | 10.34 | 0.757 | 17.185*** |
| Seafood, meat and poultry | 100g/3-day | 2.702 | 4.067 | -1.366 | -61.047*** |
| Milk, egg and nut | 100g/3-day | 0.845 | 1.602 | -0.757 | -58.694*** |
| Sugar and salt | 100g/3-day | 0.327 | 0.331 | -0.00300 | -1.499 |
| Oil | 100g/3-day | 1.118 | 1.179 | -0.0610 | -9.109*** |
| Air conditioner | - | 0.0950 | 0.240 | -0.146 | -62.672*** |
| Fan | - | 0.710 | 0.843 | -0.134 | -45.155*** |
| Refrigerator | - | 0.300 | 0.653 | -0.353 | -110.679*** |
| Heating system | - | 0.0770 | 0.0850 | -0.00700 | -3.829*** |
|  |  | Poor | Rich | diff | t-value |
| Total consumption | 100g/3-day | 29.89 | 31.27 | -1.380 | -19.607*** |
| Energy | 100kcal/3-day | 63.49 | 64.46 | -0.961 | -7.239*** |
| Carbohydrate | 100g/3-day | 10.12 | 9.290 | 0.834 | 34.175*** |
| Protein | 100g/3-day | 1.873 | 2.033 | -0.160 | -35.395*** |
| Fat | 100g/3-day | 1.717 | 2.140 | -0.422 | -62.542*** |
| Proportion of energy from fat | - | 0.246 | 0.297 | -0.0510 | -69.185*** |
| High-fat diet | - | 0.304 | 0.485 | -0.181 | -57.192*** |
| Cereal, tuber and dried legumes | 100g/3-day | 16.21 | 14.89 | 1.323 | 30.561*** |
| Vegetable, fruit and fungus | 100g/3-day | 10.63 | 11.07 | -0.433 | -10.777*** |
| Seafood, meat and poultry | 100g/3-day | 2.307 | 3.909 | -1.601 | -79.563*** |
| Milk, egg and nut | 100g/3-day | 0.735 | 1.405 | -0.669 | -56.912*** |
| Sugar and salt | 100g/3-day | 0.313 | 0.344 | -0.0310 | -15.653*** |
| Oil | 100g/3-day | 1.069 | 1.206 | -0.137 | -22.641*** |
| Air conditioner | - | 0.0810 | 0.193 | -0.112 | -52.842*** |
| Fan | - | 0.698 | 0.799 | -0.101 | -37.596*** |
| Refrigerator | - | 0.267 | 0.539 | -0.272 | -92.506*** |
| Heating system | - | 0.0710 | 0.0890 | -0.0190 | -11.070*** |

Notes: (1) This table shows the mean value of outcome variables and four appliances between four subgroups. (2) The unit of food and macronutrients are shown in the table. (3) The mean value of appliances could be seen as the owning rate in each subgroup.

**Table A13.** Adaptation effects – fans, air conditioners, heating systems, and refrigerators

| Variables | (1)<br>Proportion<br>of energy<br>from fat | (2)<br>High-fat diet | (3)<br>Total<br>consumption | (4)<br>Energy | (5)<br>Carbohydrate | (6)<br>Protein | (7)<br>Fat |
| --- | --- | --- | --- | --- | --- | --- | --- |
| 3-day CDD | 0.2702***<br>(0.0664) | 0.8821***<br>(0.2863) | -0.6092**<br>(0.2524) | -0.4410**<br>(0.2049) | -0.7676***<br>(0.2324) | -0.7549***<br>(0.2236) | 0.8582**<br>(0.3666) |
| 3-day HDD | 0.1373***<br>(0.0472) | 0.4800**<br>(0.2343) | 1.0328***<br>(0.1671) | 0.7203***<br>(0.1457) | 0.5100***<br>(0.1618) | 0.6873***<br>(0.1707) | 1.4105***<br>(0.2555) |
| 3-day CDD * fan | -0.2040***<br>(0.0650) | -0.5073*<br>(0.2775) | 0.5849**<br>(0.2415) | 0.3848*<br>(0.2023) | 0.6568***<br>(0.2316) | 0.6562***<br>(0.2226) | -0.5498<br>(0.3517) |
| 3-day CDD * AC | 0.0349<br>(0.0474) | 0.1764<br>(0.2313) | -0.1895<br>(0.1504) | -0.2841**<br>(0.1374) | -0.4216**<br>(0.1715) | -0.2978*<br>(0.1544) | -0.4513**<br>(0.2236) |
| 3-day HDD * heat | -0.0913*<br>(0.0525) | -0.2912<br>(0.2590) | -1.2467***<br>(0.1905) | -0.9280***<br>(0.1655) | -0.8342***<br>(0.1821) | -0.7949***<br>(0.1905) | -1.3885***<br>(0.2825) |
| 3-day CDD * ref | -0.0187<br>(0.0383) | -0.0732<br>(0.1873) | 0.2296*<br>(0.1247) | 0.0015<br>(0.1108) | 0.0576<br>(0.1345) | -0.2555**<br>(0.1190) | -0.2013<br>(0.1898) |
| 3-day HDD * ref | -0.0315<br>(0.0311) | -0.0541<br>(0.1456) | 0.2769**<br>(0.1177) | 0.2959***<br>(0.0915) | 0.3920***<br>(0.1004) | 0.2134**<br>(0.1077) | 0.2108<br>(0.1599) |
| Observations | 85,644 | 85,644 | 84,604 | 85,644 | 87,103 | 87,084 | 87,075 |
| R-squared | 0.5460 | 0.4425 | 0.4804 | 0.5217 | 0.5859 | 0.4952 | 0.4905 |
| Dep. Var. Mean | 0.272 | 0.395 | 30.58 | 63.98 | 9.705 | 1.953 | 1.929 |
| Individual FE | YES | YES | YES | YES | YES | YES | YES |
| Year-month FE | YES | YES | YES | YES | YES | YES | YES |
| Province-year FE | YES | YES | YES | YES | YES | YES | YES |
| Province-month FE | YES | YES | YES | YES | YES | YES | YES |

**Table A14.** Balance of predetermined characteristics on temperature exposure

|  | (1)<br>Urban | (2)<br>Male | (3)<br>Age | (4)<br>Above-median income |
| --- | --- | --- | --- | --- |
| Panel A – Without individual fixed effects |  |  |  |  |
| 3-day CDD | −0.0100***<br>(0.0011) | −0.0007<br>(0.0013) | −0.1547***<br>(0.0561) | −0.0038***<br>(0.0012) |
| 3-day HDD | 0.0043***<br>(0.0008) | −0.0002<br>(0.0009) | −0.0111<br>(0.0342) | −0.0035***<br>(0.0008) |
| Joint F-test (all characteristics jointly predict CDD): 15.36 (p = 0.000) |  |  |  |  |
| Joint F-test (all characteristics jointly predict HDD): 16.95 (p = 0.000) |  |  |  |  |
| Panel B – With individual fixed effects |  |  |  |  |
| 3-day CDD |  |  | −0.0011<br>(0.0007) | 0.0042***<br>(0.0012) |
| 3-day HDD |  |  | 0.0014***<br>(0.0005) | −0.0044***<br>(0.0008) |
| Joint F-test (all characteristics jointly predict CDD): 9.25 (p = 0.000) |  |  |  |  |
| Joint F-test (all characteristics jointly predict HDD): 17.79 (p = 0.000) |  |  |  |  |

*Notes:* (1) This table shows the balance tests of predetermined characteristics on temperature exposure. Each column reports the coefficients from a separate regression of the indicated characteristic on the 3-day CDD and HDD, together with the baseline weather controls (humidity, precipitation, wind, and sunshine). (2) In panel A, year-month fixed effects, province-year fixed effects, and province-month fixed effects are involved; in panel B, we add individual fixed effects, which absorb the time-invariant characteristics (rural status and sex). (3) Standard errors shown in parentheses are clustered at the individual level. (4) The joint F-test rows report, within each panel, the F statistic and p-value from a regression of the 3-day CDD or HDD on all the characteristics in the panel jointly (urban, male, age, and above-median income in panel A; age and above-median income in panel B), with the same fixed effects as that panel. (5) \* p < 0.1, \*\* p < 0.05, \*\*\* p < 0.01.

**Table A15.** Baseline dietary effects with additional controls for income and age

|  | (1) | (2) | (3) | (4) |
| --- | --- | --- | --- | --- |
|  | Baseline | +Income | +Age | +Both |
| Panel A – Proportion of energy from fat |  |  |  |  |
| 3-day CDD | 0.0716**<br>(0.0283) | 0.0715**<br>(0.0283) | 0.0715**<br>(0.0283) | 0.0714**<br>(0.0283) |
| 3-day HDD | 0.0376*<br>(0.0209) | 0.0377*<br>(0.0209) | 0.0377*<br>(0.0209) | 0.0379*<br>(0.0209) |
| Panel B – High-fat diet |  |  |  |  |
| 3-day CDD | 0.3842***<br>(0.1368) | 0.3824***<br>(0.1369) | 0.3835***<br>(0.1368) | 0.3817***<br>(0.1369) |
| 3-day HDD | 0.1881*<br>(0.1024) | 0.1907*<br>(0.1024) | 0.1896*<br>(0.1025) | 0.1922*<br>(0.1025) |
| Income control | No | Yes | No | Yes |
| Age control | No | No | Yes | Yes |
| Observations | 85,235 | 85,235 | 85,235 | 85,235 |

*Notes:* (1) This table shows the baseline effects with additional controls for income and age. The dependent variables are the proportion of energy from fat in panel A and the high-fat diet index in panel B. (2) The income control is per capita household income deflated to constant prices; the age control is age in years. (3) The coefficients and standard errors are multiplied by 100; the estimates are therefore interpretable as percentage-point changes per degree-day. (4) Individual fixed effects, year-month fixed effects, province-year fixed effects, and province-month fixed effects are involved in all regressions, together with the baseline weather controls (humidity, precipitation, wind, and sunshine). (5) Standard errors shown in parentheses are clustered at the individual level. (6) \* $p < 0.1$ , \*\* $p < 0.05$ , \*\*\* $p < 0.01$ . (7) All columns are estimated on the common sample with non-missing income and age, which is slightly smaller than the estimation sample of the main specification (86,147); column (1) therefore differs slightly from the baseline estimates in Table A3 (0.0807 for the CDD coefficient in panel A and 0.4039 in panel B).

**Table A16.** Temperature effects on dietary composition, by occupational group

| Variables | (1)<br>Proportion of energy from fat | (2)<br>High-fat diet |
| --- | --- | --- |
| 3-day CDD * White-collar | 0.0725<br>(0.0781) | 0.1579<br>(0.3920) |
| 3-day CDD * Physical labor | 0.0635*<br>(0.0349) | 0.3380**<br>(0.1665) |
| 3-day CDD * Other | 0.0913***<br>(0.0319) | 0.4626***<br>(0.1537) |
| 3-day HDD * White-collar | 0.0057<br>(0.0356) | 0.1114<br>(0.1776) |
| 3-day HDD * Physical labor | 0.0790***<br>(0.0298) | 0.3206**<br>(0.1354) |
| 3-day HDD * Other | 0.0237<br>(0.0248) | 0.1474<br>(0.1291) |
| Wald: Physical = White-collar, CDD (p) | 0.910 | 0.648 |
| Wald: Physical = White-collar, HDD (p) | 0.079 | 0.291 |
| Individual / wave-month / province FE | Yes | Yes |
| Observations | 86,147 | 86,147 |

*Notes:* (1) This table shows the temperature effects on dietary composition by occupational group. Each column is a single regression in which the 3-day CDD and HDD are interacted with the occupational groups, and the coefficients are the group-specific temperature slopes. (2) Physical labor comprises farmers and skilled and unskilled manual workers (n=39,454); white-collar comprises professionals, administrators, and office staff (n=8,225); “Other” collects the remaining occupations and respondents not reporting an occupation (n=55,263). The group counts refer to the full sample before singleton observations are removed. (3) The coefficients and standard errors are multiplied by 100; the estimates are therefore interpretable as percentage-point changes per degree-day. (4) The Wald rows report the p-values from tests of the null hypothesis that the physical-labor and white-collar temperature coefficients are equal. (5) Individual fixed effects, year-month fixed effects, province-year fixed effects, and province-month fixed effects are involved in all regressions, together with the baseline weather controls (humidity, precipitation, wind, and sunshine). (6) Standard errors shown in parentheses are clustered at the individual level. (7) \* p < 0.1, \*\* p < 0.05, \*\*\* p < 0.01.

## Footnotes

1 After partialling out all four sets of fixed effects and controls, the residual standard deviation of three-day CDD is 0.81 degree-days and of three-day HDD is 1.23 degree days, or 33.37% and 42.23% of the raw standard deviations, leaving substantial short-run variation that annual or monthly panels cannot exploit.

2 For more details on the literature focusing on temperature-induced reductions in crop output and agricultural TFP, see Table A1.

3 Although the CHNS has continued beyond 2011, its detailed food consumption and nutrient intake data are only available up to 2011, with no subsequent updates. Therefore, our analysis is limited to this period. We provide more details on data construction in Appendix B1. Food Consumption. Considering that off-comfort temperatures have become more frequent in the past fifteen years under climate change, the aggregate dietary burden implied by our per-degree-day estimates is likely larger today than during the sample period.

4 The same weather data and interpolation procedure are used in Ma et al. (2026). In the full national dataset, an 80 km radius leaves 98.7% of counties with at least one contributing station and draws on 10.7 stations per county on average.

5 These two cutoffs are also applied in the *Uniform standard for the design of civil buildings (GB 50352-2019)*, which suggests that the number of days with annual average daily temperatures below 5°C and above 25 °C can be used as an auxiliary indicator when dividing the temperature zones (for details and the original version in Chinese, see Figure A3). For example, if the average temperature for three consecutive days is 23°C, 25°C, and 27°C, the CDD for these three days is 0°C, 0°C, and 2°C, and the 3-day CDD for these three days is 2°C.

6 Figure A4 further demonstrates that when the daily average temperature reaches 5°C or 25°C, the corresponding minimum or maximum daily temperatures are already significantly lower or higher, respectively, indicating actual exposure to relatively cold or hot conditions even at these seemingly moderate average temperature levels. This supports the validity of our temperature thresholds in capturing meaningful cold and heat exposure episodes for our empirical analysis.

7 These three sets of fixed effects operate along distinct dimensions and are not collinear: year-month fixed effects absorb nationwide temporal variation common to all provinces, province-year fixed effects absorb province-specific annual variation, and province-month fixed effects absorb province-specific seasonal variation. Together, they absorb nationwide, province-by-year, and province-by-season variation without redundancy, leaving within-province-month deviations in the timing and weather of interviews as the identifying variation.

8 Empirical analysis of EU food supply chains documents transmission lags of approximately six months from agricultural production to food manufacturers, and a further six months from manufacturers to retail consumer (Commission of the European Communities, 2009). Given that China’s food markets during our sample period were considerably more fragmented and less integrated than those of the EU, transmission lags in China are likely at least as long as, and plausibly longer than, those documented for the EU.

9 The State Council’s 2019 white paper on food security explicitly states that emergency grain reserves are maintained in areas prone to price fluctuation and that the reserve network is activated in response to natural disasters and other transitory disruptions (State Council Information Office, 2019).

10 These arguments, like the growing-season controls in Appendix E, address supply operating through crop yields. They do not cover a distinct channel raised by perishables such as vegetables, seafood, and fruit, whose local availability can respond to contemporaneous heat within days. Two features of the results speak against this channel. First, a heat-induced shortfall in perishables would predict lower vegetable and fruit consumption on hot days, whereas Table A8 shows that vegetables, fruits, and fungi rise significantly under heat; the decline is concentrated in seafood, meat, and poultry, the foods for which heat-induced appetite suppression is best documented. Second, the response is not confined to the heat margin: fat intake rises under cold in the same specification, where no spoilage mechanism operates. A supply channel for meat and seafood alone cannot be excluded without price data. Community-level prices would permit a sharper test, and we note their absence as a limitation.

11 To illustrate the economic magnitude, a three-day heat wave with daily temperatures of 30°C on each day (cumulative CDD of 15°C) raises the proportion of dietary energy from fat by 1.215 percentage points (4.47% of the sample mean, 10.6% of the standard deviation) and the probability of a high-fat diet by 6.06 percentage points (15.3% of the mean, 12.4% of the standard deviation). Three consecutive days at a daily temperature of 0°C (HDD of 15°C) increase the fat share by 0.570 percentage points (2.10% of the mean, 4.96% of the standard deviation) and the probability of a high-fat diet by 2.925 percentage points (7.41% of the mean, 5.98% of the standard deviation).

12 Air conditioner is widely seen as the main appliance for mitigating the effects of high temperatures (Davis and Gertler, 2015; Barreca et al., 2016; Mullins and White, 2019; Randazzo et al., 2023). However, in our dataset, the ownership of air conditioners is only 13.7%. By contrast, about 75% of individuals have at least one fan in their household (Table A2). These ratios appear reasonable given that fans are less expensive than air conditioners. Thus, solely considering air conditioners would underestimate the overall effect. Therefore, we consider the roles of both fans and air conditioners.

13 We caution that appliance ownership is correlated with household wealth and urban residence, so the interaction estimates should not be interpreted as the causal effect of the technology itself, but rather as indicative of one pathway through which socioeconomic advantages may translate into dietary resilience.

14 The sample period (1991 to 2011) predates the widespread adoption of smartphones and online food-delivery platforms, so away-from-home consumption in our data reflects in-person dining rather than delivery services. In China, online food-delivery platforms expanded across cities only from 2011 onward, with the median city not reaching coverage until 2014 (Fang et al., 2026).

